# Effect of a digitally-enabled, stage-based intervention on maternal and child health: a community-based, cluster-randomized controlled trial from rural China

**DOI:** 10.64898/2026.03.27.26349570

**Authors:** Yunwei Chen, Yuju Wu, Ann M. Weber, Alexis Medina, Yian Guo, Siva Balakrishnan, Hanwen Zhang, Scott Rozelle, Huan Zhou, Gary L. Darmstadt, Sean Sylvia

## Abstract

**Background:** Comprehensive and responsive interventions are increasingly advocated to address the diverse and evolving health challenges faced by mothers and children during the first 1,000 days of life. However, evidence remains limited on how such interventions can be operationalized in low-resource settings without overstretching frontline health workers. We developed the Healthy Future program, a digitally-enabled community health worker (CHW) intervention that integrates a comprehensive and stage-based maternal and child health curriculum with digital infrastructure to deliver developmentally appropriate support through home visits. We evaluated the effects of the program on multidimensional outcomes, including caregiving practice, child health and nutrition, and caregiver well-being in rural China.

**Methods and Findings:** We conducted a cluster-randomized controlled trial across 119 rural townships in Sichuan Province, China, with 40 randomly assigned to receive the program delivered by township-level CHWs and 79 serving as controls. The trial enrolled 1,306 families with pregnant women or infants younger than six months and implemented the intervention for 12 months from August 2021 to September 2022. Among intervention families, 82% participated in at least one home visit and participating families received 10.5 home visits on average, with 4856 visits delivered in total. Follow-up data were collected from 1,149 families (88%). The main analysis used an intent-to-treat approach. For the three primary outcomes, the intervention did not significantly improve hemoglobin concentration or exclusive breastfeeding, although dietary diversity increased modestly by 0.207 food groups (95% CI: 0.015 to 0.399; p=0.035). Among secondary outcomes, the intervention reduced the occurrence of child illness during the previous two weeks by 8.2 percentage points (14.0 to 2.4; p=0.006) and improved breastfeeding initiation practices, including ever breastfeeding (3.2 percentage points; 1.1 to 5.3; p=0.002) and colostrum feeding (4.4 percentage points; 0.1 to 8.6; p=0.044). The intervention also improved caregiving knowledge index (0.189 standard deviations; 0.084 to 0.294; p<0.001), perceived social support (0.208 points; 0.026 to 0.389; p=0.028), and caregiver mental health, including reductions in symptoms of depression (6.3 percentage points; 10.8 to 1.8; p=0.006), anxiety (4.9 percentage points; 8.9 to 0.9; p=0.017), and stress (4.2 percentage points; 8.0 to 0.3; p=0.034). Supplementary domain-level summary analyses aggregating related outcomes within each domain suggested coherent improvements in caregiving knowledge, feeding practices, perceived social support, and caregiver mental health. The key limitation was that COVID-19 lockdowns and quarantines necessitated the consolidation of two planned follow-up surveys into one extended follow-up survey, creating challenges for evaluating some age-specific outcomes.

**Conclusions:** The findings suggest that a digitally-enabled CHW program can deliver a complex first-1000-days intervention in low-resource settings. The intervention generated coherent improvements in some secondary maternal and caregiving outcomes, although effects on nutrition-related primary outcomes were limited. With growing advocacy for comprehensive interventions to holistically address the diverse and evolving health needs of mothers and children during critical developmental periods, this study provides lessons for how such interventions can be designed and implemented in ways that ensure comprehensive, timely support for families without overburdening frontline health workers.

**Registration:** ISRCTN16800789

## Introduction

Evidence is irrefutable that the first 1,000 days of a child’s life, from conception to their second birthday, are critical for both the immediate and long-term health, development, and well-being of children and their mothers (*1–3*). Yet mothers and children, especially those in resource-limited communities, often confront diverse and evolving health challenges throughout this period, underscoring the need for comprehensive and responsive interventions (*4*, *5*). There has been growing global advocacy for integrated approaches to address the myriad challenges families face in achieving optimal maternal and child health during this sensitive period (*6–8*). The World Health Organization (WHO)’s Nurturing Care Framework exemplifies this approach, emphasizing integrated support across health, nutrition, responsive caregiving, early learning opportunities, and safety for children and mothers (*9*). While conceptually compelling, translating such comprehensive frameworks into effective, scalable programs remains challenging (*10*). Delivering multi-component interventions places substantial demands on health systems, particularly frontline health workers. Key obstacles include, for example, logistical complexities, limited resources, and high workloads, alongside the need to tailor support to families’ changing needs over time (*11*, *12*). Evidence remains limited on how to operationalize such interventions that are comprehensive, responsive to families’ evolving needs, and feasible for large-scale delivery in low-resource settings.

This challenge is particularly salient in rural China. Despite significant progress in child survival, persistent gaps remain in key domains of maternal and child health, including suboptimal infant feeding practices (e.g., low rates of exclusive breastfeeding among newborns and limited dietary diversity among young children) (*13*, *14*), widespread micronutrient deficiencies among mothers and children (*15*, *16*), and growing concerns about perinatal mental health among mothers in rural communities (*17*). These challenges do not occur in isolation; rather, they emerge and evolve across pregnancy and early childhood (*18*). Together, they demonstrate the need for interventions that are not only comprehensive, but also responsive to the timing and context of families’ needs.

Community-based interventions delivered by paraprofessional community health workers (CHWs) have emerged as a cornerstone delivery model for health programs in resource-limited settings (*19*, *20*). CHW-led home visiting programs have demonstrated effectiveness in extending health services to underserved populations and providing support to mothers and children, particularly in remote, rural locations (*21*, *22*). However, delivering comprehensive, multi-component interventions through CHWs presents substantial implementation challenges. As the program scope expands, CHWs face increasing cognitive and logistical burdens to maintain fidelity and consistency (*12*). Without adequate support systems, these demands can compromise intervention quality, contribute to worker overload, and limit scalability. Critically, when information is poorly timed or insufficiently tailored, families may experience information overload or disengagement, further undermining effectiveness (*11*). These realities highlight the need for innovative, community-based solutions that can respond to the diverse and evolving needs of mothers and children, while preserving the fidelity and feasibility required for large-scale implementation.

Advances in digital health technologies offer a potential pathway to overcome these constraints (*23–25*). Digital tools can support CHWs in delivering complex interventions by structuring content delivery, automating decision support, and aligning intervention components with families’ needs (*26*). When integrated into program design, digital infrastructure has the potential to enable comprehensive and responsive interventions while preserving fidelity, consistency, and feasibility at scale. However, evidence remains limited on how digitally-enabled systems can be effectively integrated to operationalize comprehensive, flexible, and responsive intervention designs, and whether such integration translates into meaningful improvements across multidimensional outcomes, including caregiving behaviors and maternal and child health (*24*).

To address this gap, we designed and evaluated the *Healthy Future* project in rural China, a comprehensive, digitally-enabled, CHW-delivered intervention targeting the holistic and evolving needs of mothers and children from pregnancy through early childhood (*27*). The intervention integrates three core components. First, we developed a comprehensive and structured intervention curriculum (the *Healthy Future* curriculum, Figure 1), designed to address key domains of maternal and child health from pregnancy through 18 months of age. These domains include maternal nutrition, exclusive breastfeeding, complementary feeding, preventive health and daily care, maternal mental health, and the uptake of government health services. Recognizing that families’ needs evolve rapidly across pregnancy and early childhood, the curriculum was explicitly designed as stage-based, with content tailored to families’ developmental stages and knowledge gaps at the time of each visit. The curriculum is further organized into three-month intervals, with new content introduced early in each interval and reinforced through subsequent visits, ensuring both timely delivery and adequate reinforcement.

**Figure 1.**
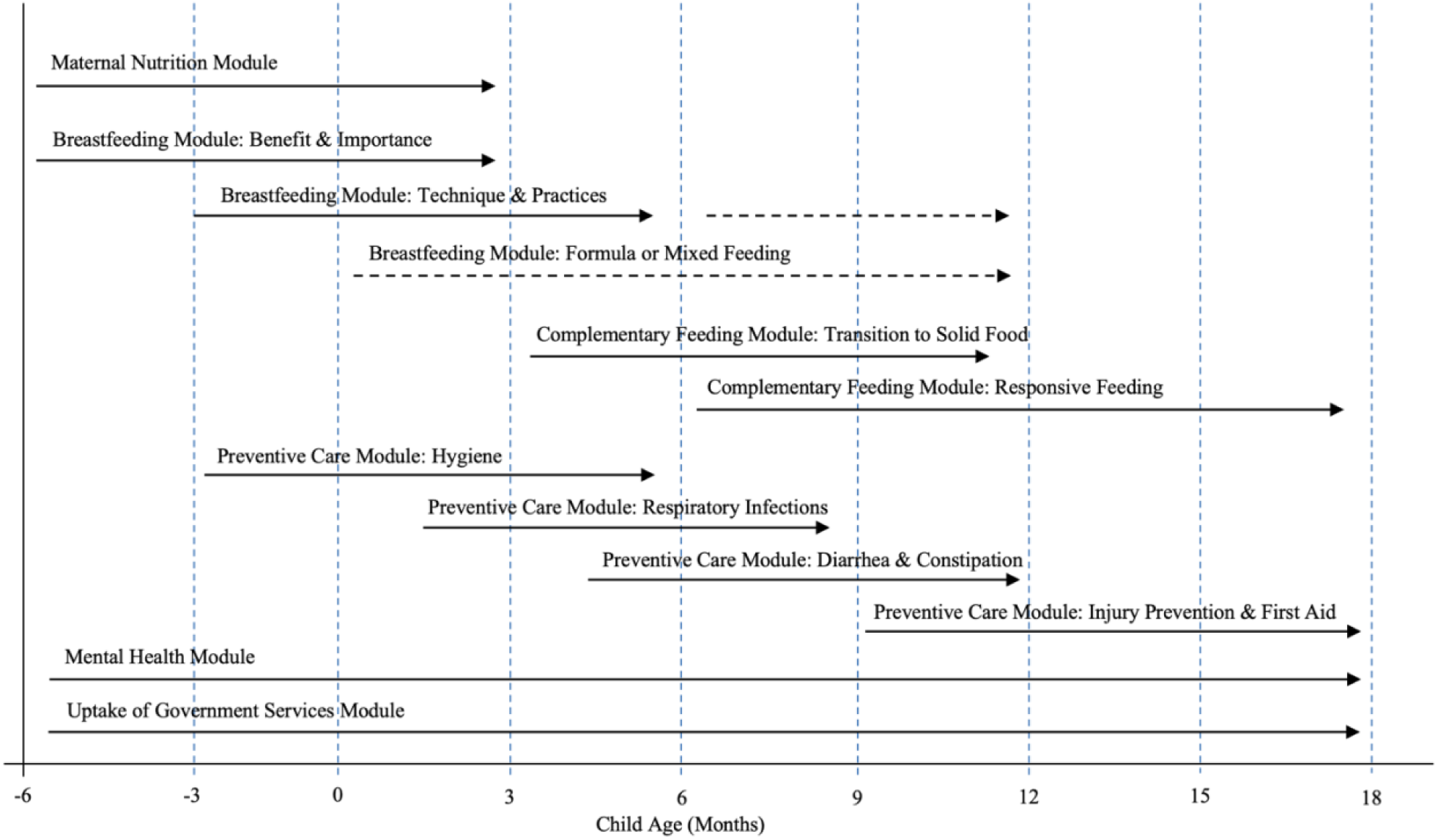
Healthy Future Curriculum. The figure illustrates the intervention curriculum design with various modules delivered at different stages of pregnancy and infancy covering pregnancy through 18 months of age.

Second, the curriculum was delivered through structured monthly home visits by CHWs, a well-established intervention model in resource-limited settings. This delivery approach allows outreach and sustained engagement with families, particularly those in underserved areas, and has the potential to be adapted to other low-resource contexts and integrated into existing community-based care infrastructures.

Third, to address the logistical challenges of delivering the right content at the right time to families at different developmental stages, we developed an integrated digital system with a tablet-based application used by CHWs (the *Healthy Future* app). Closely aligned with the curriculum design, this digital system operationalizes the intervention by automating content assignment at each visit based on families’ developmental stage and prior exposure, allowing CHWs to focus primarily on delivering content as guided by the app. By embedding curriculum content, decision support, and workflow guidance directly into the digital platform, the system aims to reduce CHWs’ cognitive and logistical burden while enabling stage-based support across diverse family trajectories and preserving consistency and delivery fidelity across CHWs and families.

Together, the *Healthy Future* program represents an effort to move beyond stand-alone interventions toward a scalable, multidimensional delivery model aimed at providing comprehensive, flexible, and timely early-life support during the critical first 1,000 days of life. This study evaluated the effects of the *Healthy Future* program on multidimensional outcomes, including caregiving behaviors, child health and nutrition, and maternal well-being using a community-based, cluster-randomized controlled trial in rural China.

## Materials and Methods

### Intervention Design

#### Curriculum development

The *Healthy Future* curriculum comprises 26 sessions across six content domains, including breastfeeding, complementary feeding, preventive health and daily care, maternal nutrition, caregiver mental health, and the uptake of government health services, designed to provide comprehensive support from the fourth month of pregnancy through 18 months after birth. A summary of the curriculum content is provided in Appendix Section S1.

The curriculum was developed by a multidisciplinary team, including researchers and practitioners with diverse expertise in economics, pediatrics, nutrition, public health, and implementation science, as well as local collaborators closely engaged with primary healthcare systems. Curriculum development was informed by established clinical and public health guidelines, local qualitative research, evidence from prior maternal and child health programs, and best practices from comparable low-resource settings. All curriculum modules underwent pilot testing prior to implementation to assess feasibility, cultural appropriateness, and acceptability in the local context. Feedback from pilot testing was incorporated to refine content and delivery before intervention initiation.

The *Healthy Future* curriculum incorporates several distinctive design features. First, it was explicitly designed as stage-based, with content timed to specific stages of pregnancy or infant age. This approach ensured that relevant information was delivered just before or when it was most needed. Home visits were scheduled monthly, with more frequent visits during the first month after childbirth to provide intensified support during particularly vulnerable periods.

Second, the curriculum was structured into intervals, typically spanning three months each. Within each interval, new content was introduced during the first month, followed by two months of content reinforcement and activities. Importantly, regardless of when families entered the program, they were exposed to the full set of content within the interval corresponding to their entry stage. For example, in the interval for pregnancy months 4-6, a pregnant woman who joined the intervention during her fifth month still began with content from the fourth month and received the full content of this interval before progressing to the next. This design ensured thorough exposure to content timed for their needs while accommodating variations in enrollment timing.

Third, the curriculum was highly scripted to facilitate delivery by minimally trained CHWs with limited prior technical knowledge, supporting consistency across home visits. Many contents were accompanied by infographic-style videos to enhance caregiver engagement and comprehension. Each visit included at least one hands-on activity to promote active participation, reinforce key messages, and support skill building. Visits followed a modular structure, with each session covering three to five core modules selected based on the child’s developmental stage. Optional modules could also be added, allowing CHWs to select relevant content based on the specific needs of each family. This modular design also enabled repetition of key messages across developmental stages and allowed schedules to be adjusted without disrupting curriculum integrity.

#### Digital integration

To address the logistical challenges of delivering timely, stage-appropriate information to families at varying developmental stages, the research team collaborated with a software company (ThoughtWorks, Inc.) to develop an integrated digital platform with a tablet-based app designed to support CHWs during intervention delivery. The platform underwent pilot testing prior to the trial and was fully operational throughout program implementation. Appendix Section S1 provides examples of app interfaces and describes its key features.

The core component of the platform is the *Healthy Future* app, a tablet-based application that supports CHWs in scheduling home visits and delivering the intervention curriculum. This app embedded a workflow algorithm closely aligned with the curriculum design and automatically assigned age-appropriate content to each family at each visit based on gestational age or child age and prior visit completion. This automation substantially reduced the logistical complexity and cognitive burden for CHWs, enabling consistent and timely delivery of content across families. The app also facilitated family-specific needs assessment and allowed CHWs to supplement core content with optional modules appropriate for each family’s circumstances. Importantly, the app supported standardized record keeping, adaptive post-visit surveys, and real-time data capture for tracking caregiver participation, knowledge acquisition, and practices following each visit.

The second component of the platform is a web-based administrative portal that allowed supervisors to remotely access tablet-collected data and monitor CHW activities. Data were also aggregated and analyzed through a dashboard that tracked the timeliness, frequency, duration, completeness, and quality of the visits. With this system, supervisors were able to quickly identify potential implementation issues, such as unusually short visits or systematic deviations in visit durations across CHWs and provide fast feedback and corrective support as needed.

Overall, this digital platform enables flexible yet standardized delivery of a complex, multi-component intervention in response to families’ diverse and evolving needs. The app is now open-source and translatable into other languages, offering an adaptable model for maternal and child health programs in other resource-limited settings globally.

Because the *Healthy Future* program consisted of health education, counseling, and supportive home visits, no major intervention-related adverse events or unintended harms were anticipated. The intervention did not involve clinical procedures, medical treatments, or the provision of medications.

### Experimental Design

#### Study setting

A study protocol has been published detailing the study design, rationale, determination of sample size, and evaluation methods (*27*). Briefly, to evaluate the effect of the *Healthy Future* program, we conducted a parallel cluster-randomized controlled trial in Sichuan Province, China, across 119 rural townships (clusters) from four counties previously designated as national poverty-stricken counties. The sampling frame included all rural townships within these four counties with a population of at least 10,000, ensuring a representative rural cohort and sufficient statistical power.

#### Randomization and masking

We conducted a stratified cluster-randomized controlled trial in which rural townships were assigned to either the intervention or control arm. Randomization at the township level was used to minimize contamination, as CHWs delivered the intervention at the township level. Townships were stratified by county (four strata) and randomly allocated to study arms using Stata/SE 15.0 with the *randtreat* command (*28*). Of the 119 townships, 40 (ten per county) were randomly assigned to receive the *Healthy Future* program, while the remaining 79 townships served as controls and received no intervention. At the same time, both treatment and control groups received routine maternal and child health services through local township health centers.

Randomization and data analyses were conducted by individuals not involved in the program’s implementation. Families in the control group participated only in household surveys without learning about the broader program messages. Masking families and CHWs in the intervention group was not possible. Enumerators for the household surveys, recruited separately before each survey, were unaware of the study design, treatment assignment, and evaluation hypotheses. CHWs received training exclusively on intervention delivery and were blinded to the trial design or evaluation objectives.

#### Participant recruitment

Families with pregnant women in their second or third trimesters and families with infants aged 0-6 months were enrolled at baseline, before program implementation, across all 119 townships. The trial was designed to be implemented for 12 months, covering pregnancy through 18 months of age. Recruitment was conducted through two channels: First, we collaborated with local township health centers that maintained lists used by local governments to manage care for pregnant women and newborns within their catchment areas. Second, we obtained lists of pregnant women from county-level hospitals to identify pregnant women who might bypass local township health centers for prenatal care. Additionally, survey teams randomly checked local communities to ensure that no pregnant women or infants were omitted from the primary sources.

Eligible participants were residents of the 119 study townships who were either pregnant women in the second trimester or later, or caregivers of infants aged 0-6 months, and who were willing and able to provide informed oral consent and participate in the survey. The study included all eligible families in study townships that provided informed oral consent. In townships with more than 30 eligible families, a maximum of 30 families were randomly enrolled. Participants in 40 treatment townships were eligible to receive the *Healthy Future* intervention.

#### CHW recruitment, training, and supervision

Each intervention township employed one CHW from the local community to deliver the intervention through home visits, with a total of 40 CHWs involved in the trial. CHWs were recruited by local township health centers and selected through interviews conducted by the study team. Eligible criteria for CHWs included being female, having completed at least middle school education, and being a long-term township resident. CHWs were employed for the study and compensated based on the number of families they managed.

All CHWs received five-day baseline training prior to intervention launch, as well as two-day refresher training approximately six months later. Training covered curriculum content delivery, interpersonal communication skills, use of the *Health Future* app, and role-play exercises emphasizing effective interactions with families. The training emphasized interactive dialogues over didactic information delivery. Multiple role-play sessions were also incorporated to allow CHWs to practice conducting home visits, familiarize themselves with the curriculum content and delivery formats, navigate the tablet-based app, and receive structured feedback on communication style and visit execution.

At intervention initiation, members of the implementation team accompanied CHWs during initial home visits to introduce the program and enroll families to participate in the intervention. Thereafter, CHWs conducted monthly home visits for approximately 12 months, with more frequent visits during the first month following childbirth. Ongoing supervision included monthly check-ins with CHWs to provide support and address implementation challenges. Treatment households were also contacted periodically to assess satisfaction with CHW interactions and home visits. In addition, a trial monitoring team conducted random monthly phone calls with three participant families per CHW to assess satisfaction and visit quality. The team also conducted at least one in-person observation of home visits with each CHW every three months to ensure adherence to study protocols.

#### Implementation

The study team was established in 2018 and focused initially on developing a comprehensive and stage-based curriculum (i.e., the *Healthy Future* curriculum) targeting the holistic and evolving health needs of pregnant women and young children. In early 2020, efforts shifted to developing the mHealth system with a tablet-based application (i.e., the *Healthy Future* app) to facilitate intervention delivery. Between July and August 2021, we completed the baseline survey, recruited and trained CHWs, and enrolled participants. The intervention was officially launched in August 2021 and implemented until September 2022.

We originally planned two follow-up surveys at 6 and 12 months after enrollment. However, frequent COVID-19 lockdowns and quarantines during the study period substantially disrupted data collection. To preserve follow-up and intervention continuity as much as possible, we modified the plan to prioritize achieving a high follow-up rate with a single extended follow-up survey administered in person and by telephone in two phases: March to May 2022 and August to October 2022. Telephone follow-ups mitigated significant sample loss due to family refusals and migration, particularly during COVID-19 lockdowns that exacerbated concerns about disease transmission. However, telephone surveys, constrained by the practical limitations of time, were necessarily shorter and collected less information than in-person surveys.

Importantly, COVID-19 restrictions primarily affected data collection rather than program implementation. Data collection relied on large enumerator teams that were subject to travel restrictions and lockdowns, leading to delays and increased costs. In contrast, CHWs’ engagement with families was more flexible and could be readily adjusted. Missed visits did not disrupt subsequent intervention delivery, as the stage-based, interval-structured design allowed content to realign with families’ current developmental stages.

#### Data collection

Data for the impact evaluation were collected from three primary sources. First, two waves of household surveys, including baseline and follow-up, were conducted to gather comprehensive information about families, caregivers, and children. Surveys were administered using tablet-based Survey Solutions data entry software, with questionnaires programmed with built-in range checks, skip logic, and internal consistency checks to enhance data quality. Enumerators were recruited prior to each survey wave and received two-day training in survey protocols and data collection procedures. In addition to questionnaire-based data, trained nurses collected anthropometric measurements and hemoglobin concentrations for each child in each household using standardized protocols.

A typical in-person household survey lasted approximately two hours. A small subset of follow-up surveys was also conducted by telephone, to mitigate sample loss during periods of heightened concern about COVID-19 transmission. These telephone surveys were implemented during the later stages of follow-up, were necessarily shorter (approximately 30 minutes), and collected a reduced set of outcome measures compared to in-person surveys.

In addition, demographic and background information on CHWs was collected electronically on the first day of baseline CHW training. Finally, the Healthy Future app captured extensive program process data, including the timestamps and duration of each visit, modules delivered, reasons for incomplete visits, and adaptive survey responses collected following each visit.

#### Outcomes

In accordance with the pre-specified study protocol, three primary outcomes were defined. These included: (1) hemoglobin concentration among children older than six weeks, measured using the HemoCue Hb 201+ finger-prick system; (2) exclusive breastfeeding among infants younger than six months, defined as the infant having received only breast milk in the previous 24 hours with no other liquids or foods; and (3) dietary diversity among children older than six months, defined as the number of food groups consumed by the child in the previous 24 hours.

Following WHO guidelines, the two feeding measures were measured using standardized 24-hour dietary recall surveys that recorded all foods and liquids the child consumed on the day preceding the survey (*29*). Hemoglobin concentrations were measured by trained nurses during household surveys using the HemoCue Hb 201+ finger-prick system (HemoCue, Ängelholm, Sweden), contingent on caregiver consent. We note that hemoglobin measurements had lower completion rates than questionnaire-based outcomes, largely due to caregiver refusal or difficulties conducting the finger-prick procedure among young infants.

Additionally, we present treatment effects on a comprehensive set of pre-specified secondary outcomes, reflecting the integrated content domains covered by the *Healthy Future* curriculum (*27*). These secondary outcomes span child growth and illness indicators (*30*), maternal and caregiver mental health (*31*, *32*), infant and young child feeding practices (*29*), caregiver hygiene practices, mothers’ prenatal care, and intermediate outcomes, including caregiver knowledge and perceived social support (*33*). Whenever possible, gold standard or locally-validated measures were used, such as the Edinburgh Postnatal Depression Scale (*31*), Depression, Anxiety, and Stress Scales – 21 Items (*32*), and the Multidimensional Scale of Perceived Social Support (*33*). Table A in Appendix S2 outlines the full list of these outcome measures and summarizes deviations from the original study protocol.

### Statistical Analysis

#### Main evaluation specification

Our main evaluation employed intent-to-treat (ITT) analysis to estimate the effects of offering the program to eligible families. We estimated the following econometric specification:

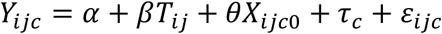

*Y*_*ijc*_ is the outcome of interest for individual *i* (child or caregiver) in township *j* of county *c* measured at follow-up; *T*_*ij*_ is an indicator for whether the individual *i* in township *j* was assigned to the treatment arm; *τ*_*c*_ denotes randomization strata (i.e., county *c*) fixed effects; *X*_*ijc*0_ is a vector of baseline control variables, capturing child, caregiver, and household characteristics measured at baseline, selected using a machine-learning post-double-selection lasso procedure, which enhances statistical power for detecting true effects while preventing over-selection of spurious covariates (*34–36*). Standard errors were clustered at the township level using the cluster-corrected Huber-White estimator. The coefficient of interest, *β*, represents the ITT effect of offering the program. All data analyses were conducted using State/MP 19.0.

The post-double-selection lasso procedure has become widely adopted in applied econometric research for estimating treatment effects in experiment settings (*37*, *38*). The motivation for using the post-double-selection lasso procedure is that the appropriate set of control variables is typically unknown ex ante. Even under successful randomization, failing to control for strong predictors of the outcome can substantially reduce statistical power. This method addresses this trade-off by selecting valid covariates from a high-dimensional set of baseline variables, thereby improving precision in estimating treatment effects while avoiding overfitting (*34*, *35*).

When estimating intervention effects on secondary outcomes, multiple related measures were considered within each domain. To account for multiple hypothesis testing across these outcomes, we adjusted the p-values within each domain using a false discovery rates (FDR) correction procedure, with the FDR-adjusted p-values reported (*39*, *40*). This procedure controls the expected proportion of false discoveries among rejected hypotheses, adjusting for multiple hypothesis testing across related outcomes within the same domain.

As we incorporated telephone surveys for a small subset of families at follow-up to mitigate sample loss during periods of heightened concern about COVID-19 transmission, outcome availability varied across survey models because telephone surveys were necessarily shorter and collected less information than in-person surveys. To address potential bias arising from differential completion rates across outcomes, we implemented a machine-learning lasso-based inverse probability weighting (IPW) approach (*41–43*). Specifically, we constructed weights based on the inverse of the predicted probability that a given outcome was observed, conditional on the rich set of baseline child, caregiver, and household characteristics estimated using a lasso procedure. Extreme weights were truncated to improve robustness. When an outcome had incomplete follow-up data, we reported both unweighted estimates and IPW-adjusted estimates to assess sensitivity to differential outcome completion.

#### Sensitivity analyses

We also conducted a compliance-adjusted treatment-on-the-treated (TOT) analysis using an instrumental variable (IV) approach, with assignment to an intervention township as an instrument for intervention dose, defined as the number of completed home visits prior to follow-up. While the main ITT estimates capture the overall intervention effects of offering the *Healthy Future* program, TOT estimates capture the effects of intervention exposure among participating families. We reported TOT results in Appendix Section S4 as sensitivity analyses.

### Ancillary analyses

To facilitate clear interpretation and capture overall intervention effects for each outcome domain, we also constructed domain-level summary indices that consolidate related outcome measures, including both primary and secondary outcomes, into a single index per domain, using a generalized least squares weighting procedure (*39*). The motivation is that, although a broad range of pre-specified primary and secondary outcomes were examined to reflect the integrated content areas covered by the intervention, this breadth also posed challenges for interpreting overall intervention effects across multiple outcome measures from multiple domains, especially when the magnitude or direction of effects varies across individual indicators. These domain-level indices enable a more comprehensive assessment of program effects than analyses based solely on individual outcome measures when evaluating comprehensive, multi-component interventions such as the *Healthy Future* program. The generalized least squares weighting procedure computes a weighted mean of standardized outcome variables, using weights derived from the inverse of the covariance matrix of the standardized individual indicators. Under this procedure, highly correlated indicators receive smaller weights, while less correlated outcomes receive larger weights, thereby enhancing the representativeness and informativeness of each domain-level index (*39*). Table B in Appendix S3 provides the list of primary and secondary measures used to construct the domain-level summary indices. Indices were constructed only for domains containing multiple related outcomes measures. We applied the same index construction procedure consistently across all domains to ensure the comparability of intervention effects.

### Pre-specified vs. post-hoc analyses

ITT and TOT specifications, the use of post-double-selection lasso for selecting control variables, and adjustments for multiple-inference testing and missing outcomes were all specified in the published study protocol. Domain-level summary index analyses were added post-hoc to facilitate high-level interpretation of intervention effects across multiple related outcomes within each domain using the pre-defined set of primary and secondary outcomes. We interpret these indices as complementary summaries of intervention effects rather than replacements for individual outcome analyses. Details on all statistical methods are provided in Appendix Section S3.

### Registration

The trial was registered on July 21, 2021, with reference ISRCTN16800789. The first enrollment date was July 25, 2021.

### Ethical approval and consent

The study was approved by the institutional review boards (IRBs) at Stanford University (Protocol 44312), the University of Nevada, Reno (Project 1737966-1), and Sichuan University (Protocol K2019046). All caregivers provided informed oral consent for their own participation and that of their infants. Enumerators received standardized training on the informed consent process, including explaining the study objectives, answering participants’ questions, and obtaining informed oral consent before data collection.

### Trial protocol

The study protocol was published in 2023 (*27*).

### Deviation from Protocol

Appendix Section S2 provides detailed justification for deviations from the published study protocol. First, the protocol specified two follow-up surveys; however, repeated COVID-19 lockdowns and quarantines during the survey period substantially disrupted data collection, necessitating the consolidation of the two planned follow-up surveys into one extended follow-up survey to preserve follow-up and minimize sample loss. Second, the protocol specified two intervention arms: a standard intervention arm and a family encouragement arm. Since the main objective of this paper is to describe the development of the *Healthy Future* program and to evaluate its overall effectiveness, we combined these two intervention arms and estimated the overall effects of offering the program relative to the control group. Third, we modified some pre-specified secondary outcomes to account for updates to WHO guidelines for assessing infant and young child feeding practices and to adjust for the consolidation of two follow-up surveys, which reduced the sample size for evaluating some age-specific outcomes. Fourth, we added domain-level summary indices to facilitate high-level interpretation of the program’s overall impacts across domains and to improve the interpretability of findings across multiple related outcomes that may exhibit divergent patterns. Developing domain-level indices with this approach has become increasingly common in the evaluation of multi-component, complex interventions (*44–46*). Lastly, we note that the trial was initially registered with 80 townships, including 40 treatment and 40 control townships. After the baseline survey, we found cluster sizes to be smaller than anticipated and therefore enrolled 39 additional control townships for powering up, resulting in 119 total townships (40 treatment and 79 control). This modification was made during the early stages of the trial and has been incorporated into the published protocol.

## Results

### Sample Description

The trial enrolled families at varying developmental stages at baseline, ranging from the second trimester of pregnancy through infancy younger than 6 months, with a planned implementation period of 12 months. This design allows coverage of families across the continuum from pregnancy through 18 months of child age and reflects the real-world context in which CHWs manage multiple families at different stages. Enrollment occurred between July and August 2021, prior to program implementation. A total of 1306 families from 119 townships were enrolled, including 562 families in the intervention arm and 744 families in the control arm (Figure 2). At baseline, 499 families (38%) included pregnant women, and 807 families (62%) included infants younger than 6 months. Figure A in Appendix S1 illustrates the distribution of gestational age and infant age at the time of enrollment.

**Figure 2.**
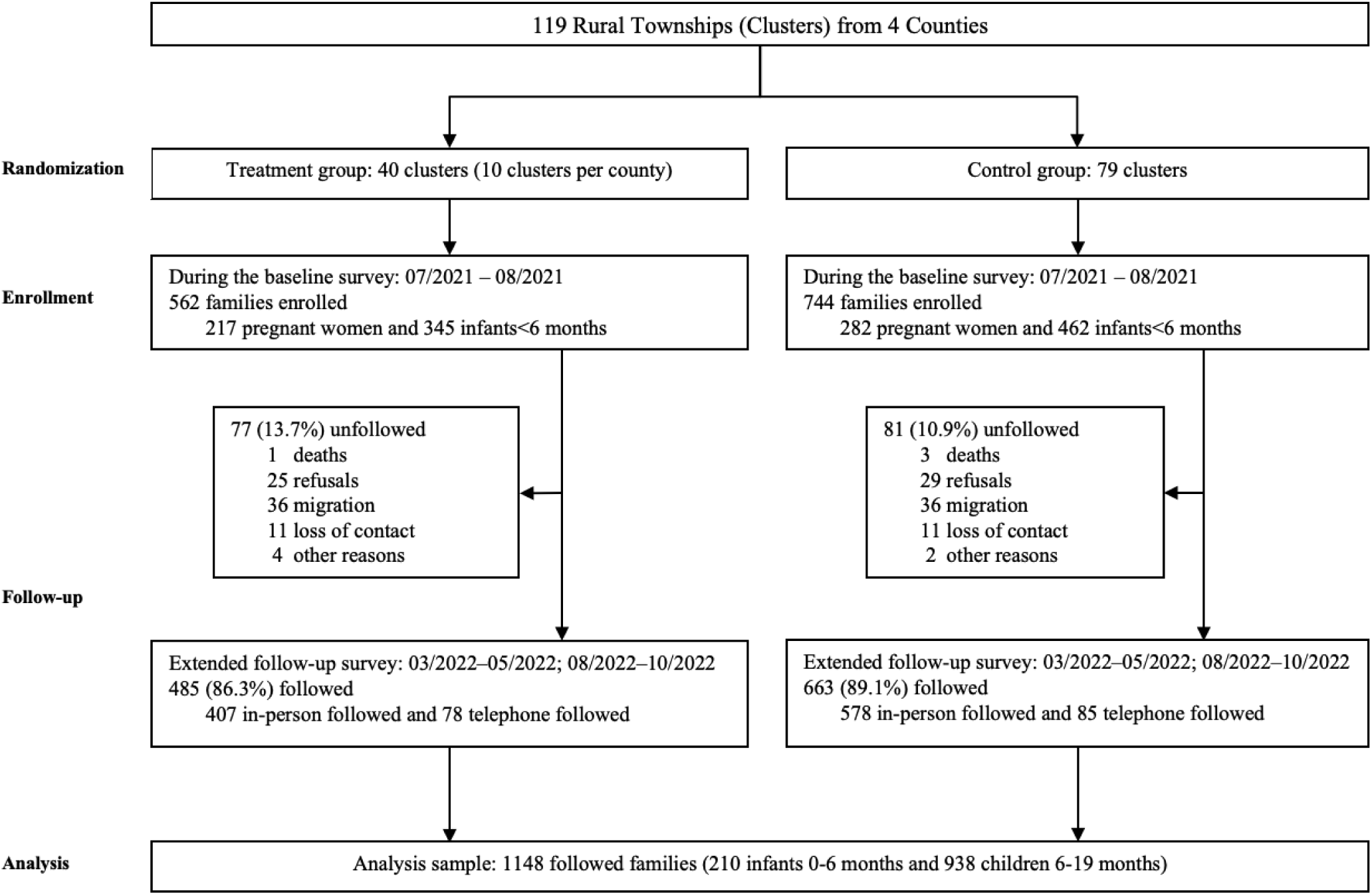
**Trial profile.**

Table 1 summarizes the baseline characteristics of children, their caregivers, and households. Among the 807 infants enrolled at baseline, only 258 (32%) were currently exclusively breastfed, indicating low adherence to recommended feeding practices for infants younger than 6 months. Only 79 (10%) of infants received exclusive breastfeeding during the first two days after birth, despite all deliveries occurring in hospitals or healthcare facilities. Hemoglobin measurements were obtained for 634 infants (79%), of whom 274 (43%) were identified as anemic (hemoglobin levels <110 g/L). Mental health assessments among primary caregivers, predominantly pregnant women or mothers of infants, revealed that 250 (19%) showed depression symptoms, 272 (21%) anxiety symptoms, and 154 (12%) elevated stress. Together, these baseline characteristics represent the multiple and concurrent health challenges faced by families during pregnancy and early infancy. Baseline characteristics were well balanced between intervention and control groups across child, caregiver, and household characteristics.

**Table 1.** Baseline characteristics.

|  | All<br>(N=1306) | Control<br>(N=744) | Treatment<br>(N=562) | P-values |
| --- | --- | --- | --- | --- |
| **Panel A. Baby Characteristics** |  |  |  |  |
| Baseline status |  |  |  |  |
| Baseline pregnant | 499 (38%) | 282 (38%) | 217 (39%) | 0.79 |
| Baseline baby | 807 (62%) | 462 (62%) | 345 (61%) |  |
| Pregnancy weeks (if unborn) | 27.1 (7.6) | 27.5 (7.5) | 26.6 (7.7) | 0.20 |
| Baby age (months, if born)* | 3.2 (1.7) | 3.1 (1.7) | 3.2 (1.7) | 0.38 |
| Baby gender (if born)* |  |  |  |  |
| Girl | 400/807 (50%) | 228/462 (49%) | 172/345 (50%) | 0.89 |
| Boy | 407/807 (50%) | 234/462 (51%) | 173/345 (50%) |  |
| Anemia (0/1)* | 274/634 (43%) | 156/362 (43%) | 118/272 (43%) | 0.94 |
| Exclusive breastfeeding (0/1)* | 258/803 (32%) | 152/461 (33%) | 106/342 (31%) | 0.55 |
| Exclusive breastfed first 2 days after birth (0/1)* | 79/805 (10%) | 43/460 (9%) | 36/345 (10%) | 0.61 |
| **Panel B. Caregiver Mental Health** |  |  |  |  |
| Depression (0/1) | 250 (19%) | 142 (19%) | 108 (20%) | 0.92 |
| Anxiety (0/1) | 272 (21%) | 155 (21%) | 117 (21%) | 0.97 |
| Stress (0/1) | 154 (12%) | 87 (12%) | 67 (12%) | 0.88 |
| **Panel C. Household Characteristics** |  |  |  |  |
| The primary caregiver was mother (0/1) | 1256 (96%) | 720 (97%) | 536 (95%) | 0.19 |
| A secondary caregiver was present (0/1) | 980 (75%) | 553 (74%) | 427 (76%) | 0.49 |
| Grandparents lived together at baseline (0/1) | 1100 (84%) | 625 (84%) | 475 (85%) | 0.74 |
| Family size (number) | 5.4 (1.2) | 5.5 (1.2) | 5.4 (1.2) | 0.30 |
| The baby had siblings (0/1) | 825 (63%) | 471 (63%) | 354 (63%) | 0.89 |
| Mother's age | 28.5 (5.0) | 28.5 (4.9) | 28.5 (5.1) | 0.82 |
| Mother's education: high school or higher (0/1) | 506 (39%) | 290 (39%) | 216 (39%) | 0.82 |

Data are n (%), mean (SD), or n/N (%). \*Data are available only for baseline babies.

A total of 40 CHWs were recruited for program implementation, with one CHW employed in each intervention township. CHWs were selected from their local communities and delivered the *Healthy Future* intervention through monthly home visits, supported by the *Healthy Future* app. CHW characteristics are summarized in Table A in Appendix S1. All CHWs were females, with a mean age of 34.6 years at program initiation. Half (52%) had completed middle school, 9 (22%) had completed high school, and 10 (25%) had attained a college or vocational college degree. All CHWs were mothers themselves, and 34 (85%) reported having two or more children. Most CHWs (25, 62%) were employed full-time for the program.

### Program Completion

Program implementation began in August 2021 and concluded in September 2022, lasting approximately 12 months. No major intervention-related adverse events or unintended harms were reported during the study period.

Because families were enrolled at varying developmental stages, from the second trimester of pregnancy through infancy younger than 6 months, the intervention was structured to span the continuum from pregnancy through 18 months of child age. As a result, families experienced different patterns of content exposure depending on their entry point into the program. For example, families enrolled during pregnancy received greater exposure to antenatal care and breastfeeding content, whereas those enrolled during infancy received more extensive coverage of complementary feeding and preventive care during the implementation period. Modules related to caregiver mental health were delivered across all stages to provide continuous support throughout pregnancy and early childhood. This variation in content exposure reflects the intended stage-based design of the intervention, in which families receive content aligned with their developmental stages and needs.

Figure 3 presents the distribution of completed home visits across developmental stages, from the fourth month of pregnancy (H4) through 18 months of child age (B18). The figure reflects the stage-based, interval-structured design of the intervention, which typically spans three months per interval with new content introduced in the first month and reinforced during the subsequent two months. To accommodate the variation in families’ entry points, families always began with the first session of the interval corresponding to their enrollment stage and were able to complete all sessions within that interval before progressing to the next. When families transitioned to a new developmental interval before completing all sessions, uncompleted sessions were skipped and subsequent visits automatically advanced to the next interval. These scheduling and content-assignment rules were embedded within the *Healthy Future* app, which automatically assigned visit content based on the timing of the home visit, the family’s current developmental stage, and prior visit completion. Overall, Figure 3 illustrates how intervention delivery was distributed across pregnancy and early childhood, aligning visit timing and content with families’ progression through early developmental stages.

**Figure 3.**
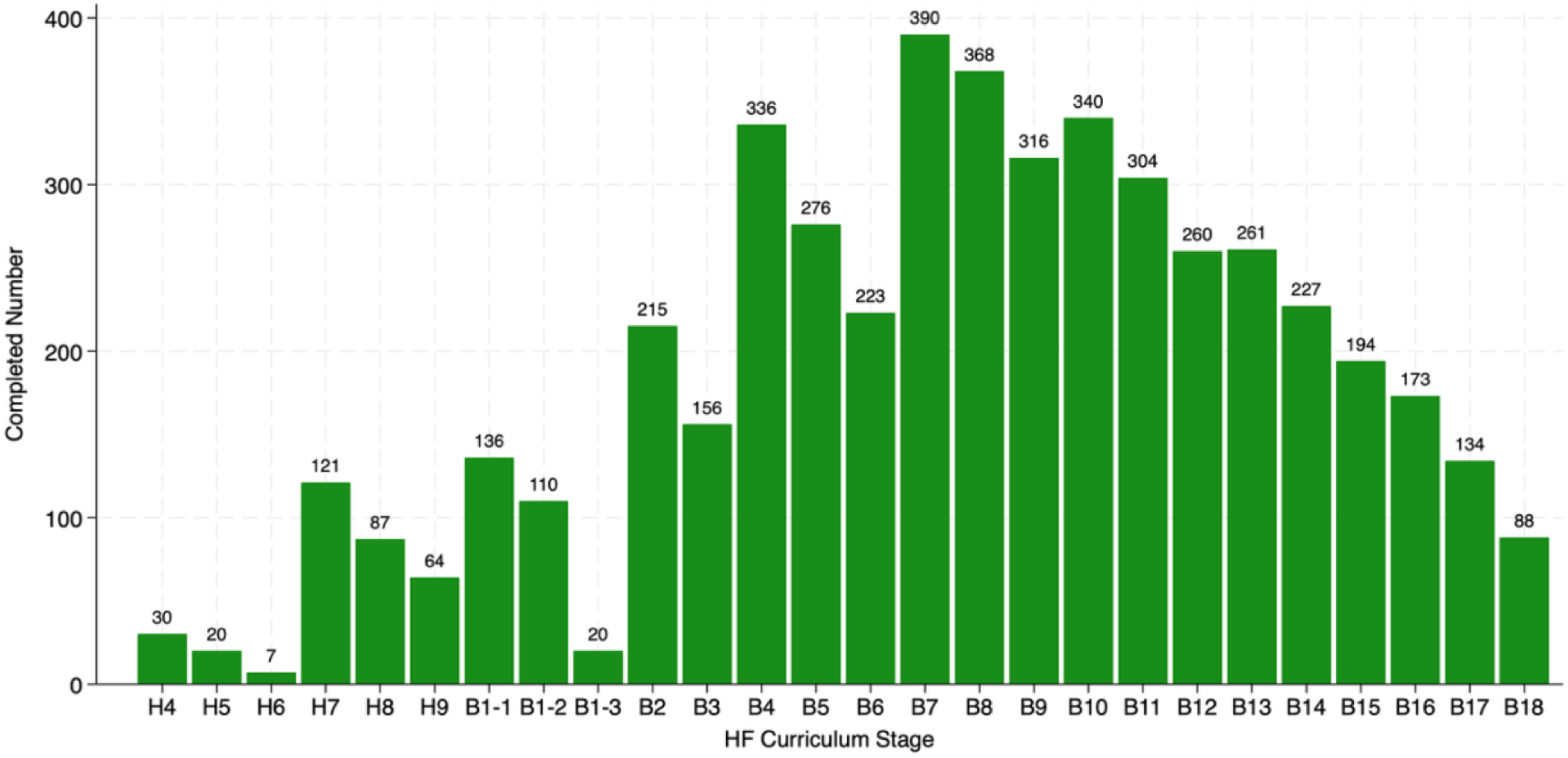
Intervention completion by developmental stages. This figure graphically illustrates the distribution of home visits across various stages of pregnancy and infancy. The Y-axis displays the number of completed home visits. The X-axis presents the stages from the fourth month of pregnancy (H4) to the eighteenth month of infancy (B18). The standard protocol calls for monthly visits but designed three home visits in the first month post-delivery, represented by B1-1, B1-2, and B1-3, to provide enhanced support immediately after birth. This figure highlights the interval-based structure of the curriculum that typically spans three months per interval. Regardless of where families entered the program, they always began with the first session of the interval corresponding to their stage, which introduced new content followed by two months of reinforcement.

Table 2 summarizes program process data captured by the *Healthy Future* app, which not only supports delivery fidelity but also enables systematic tracking of implementation. The data indicate high intervention uptake: among the 562 families in intervention townships, 461 (82%) participated in at least one home visit. Over the 12-month implementation period, a total of 4,856 home visits were completed among all participating families, with a median visit duration of 32 minutes. Participating families received an average of 10.5 home visits, with a median of 12 and a range of 1 to 16 visits. Figure C in Appendix S1 illustrates the distribution of completed home visits among participating families, showing that 75% completed at least 10 home visits. These patterns are consistent with the program design. Although the standard protocol scheduled monthly visits over 12 months, such that a typical family would be expected to receive 12 visits in total, some families received additional visits for two reasons. First, visits were more intensive during the first month following childbirth, with three visits scheduled. Second, the interval-structured curriculum allowed families who enrolled later within an interval to complete content from earlier stages of that interval.

**Table 2.** Home-visit completion statistics for the intervention families.

|  | Treated Families |
| --- | --- |
| **Panel A. Intervention take-up (N=562)** |  |
| Ever participated (0/1) | 461 (82%) |
| **Panel B. Completion, if participated (N=461)** |  |
| Number of home visits completed | 10.5 (3.9) |
| Number of breastfeeding modules completed | 7.5 (3.7) |
| Number of complementary feeding modules completed | 6.7 (3.0) |
| Number of preventive health & daily care modules completed | 6.5 (2.7) |
| Number of maternal nutrition modules completed | 2.1 (2.7) |
| Number of caregiver mental health modules completed | 5.2 (2.5) |
| Number of government health services modules completed | 3.3 (1.5) |

Data are n (%) or mean (SD).

Table 2 also reports completion across specific content domains. Breastfeeding was the most frequently delivered module, with participating families receiving an average of 7.5 sessions. This was followed by complementary feeding (6.7 sessions), preventive health and daily care (6.5 sessions), caregiver mental health (5.2 sessions), government health services (3.3 sessions), and maternal nutrition (2.1 sessions). These differences in domain-specific exposure reflect the intended stage-based design of the intervention, in which families received content aligned with their developmental stages and needs rather than equal exposure across all domains.

### Follow-up and Attrition

Overall, of the 1306 families enrolled at baseline, 1148 (88%) were successfully followed up during the extended survey period (Figure 2), including 210 infants aged 0-6 months and 938 children aged 6-19 months. The age distribution of children at the time of follow-up is shown in Figure B in Appendix S1. Among the 1148 families followed up, 985 families were surveyed in person and 164 were reached by telephone. Among the 158 families lost to follow-up, the reported reasons included death or miscarriage (4, 0.3%), family refusal (54, 4.1%), migration (72, 5.5%), loss of contact (22, 1.7%), and other reasons (6, 0.5%). Attrition rates did not differ significantly between the intervention and control groups.

### Effects on Primary Outcomes

Table 3 presents the intervention effects on three primary outcomes: hemoglobin concentration among children older than 6 weeks measured using the HemoCue Hb 201+ finger prick system; exclusive breastfeeding among infants younger than 6 months; and dietary diversity score among children older than 6 months. The intervention modestly improved dietary diversity among children aged 6–19 months, with an average increase of 0.207 food groups (95% confidence interval (CI): 0.015 to 0.399; p=0.035). No statistically significant effects were observed on hemoglobin levels or exclusive breastfeeding among infants under 6 months.

**Table 3.** Intervention effects on primary outcomes.

|  | N<br>(eligible) | N<br>(obs) | Control<br>Mean | Overall Intervention Effect |  |  |
| --- | --- | --- | --- | --- | --- | --- |
|  |  |  |  | ITT Estimate<br>(95% CI) | IPW-adjusted Estimate<br>(95% CI) | P-value |
| Hemoglobin level (g/L, 0-19 months) | 1148 | 904 | 110.803 | -0.382 (-1.964 to 1.200) | -0.346 (-1.919 to 1.228) | 0.636 |
| Exclusive breastfeeding (0/1, 0-5 months) | 210 | 210 | 0.336 | 0.057 (-0.060 to 0.174) | -- | 0.341 |
| Dietary diversity score (food groups, 6-19 months) | 938 | 938 | 4.209 | 0.207** (0.015 to 0.399) | -- | 0.035 |

All regressions included county fixed effects (randomization strata) and controlled for child-, caregiver-, and household-level covariates selected using a post- double-selection lasso procedure. Standard errors were clustered at the township level. For outcomes with incomplete follow-up data, we implemented a machine-learning lasso-based inverse probability weighting (IPW) approach and reported both intent-to-treat (ITT) estimates and IPW-adjusted estimates to assess the sensitivity of results to differential outcome completion. P-values correspond to the ITT estimates. \*\* p<0.05 \*\*\* p<0.01

Notably, the precision of estimates varied across outcomes because some primary outcomes were measured in age-specific subsamples. Exclusive breastfeeding was assessed only among infants younger than 6 months (n=210 at follow-up). This reflects disruptions to the originally planned two-wave follow-up design caused by COVID-19 field lockdowns, which necessitated consolidation into a single extended follow-up period. As a result, sample sizes for outcomes defined over narrow age windows, such as exclusive breastfeeding, were smaller than anticipated. In contrast, a larger share of children had aged beyond 6 months by follow-up, yielding greater statistical power for outcomes measured among older children, such as dietary diversity.

### Effects on Secondary Health Outcomes

Table 4 presents the intervention effects on secondary health outcomes among children, mothers, and primary caregivers. The intervention significantly reduced the incidence of any reported child illness in the past two weeks by 8.2 percentage points (CI: 14.0 to 2.4; p=0.006). No statistically significant effects were observed on child nutritional status indicators at the time of follow-up, consistent with the expectation that changes in anthropometric and related nutritional outcomes often require longer periods to emerge.

**Table 4.** Intervention effects on secondary health outcomes.

|  | N<br>(eligible) | N<br>(obs) | Control<br>Mean | Overall Intervention Effect |  |  |  |
| --- | --- | --- | --- | --- | --- | --- | --- |
|  |  |  |  | ITT Estimate<br>(95% CI) | IPW-adjusted Estimate<br>(95% CI) | P-value | FDR-adjusted<br>P-value |
| Child Health (Children) |  |  |  |  |  |  |  |
| Anemia (0/1) | 1148 | 904 | 0.421 | 0.040 (-0.026 to 0.105) | 0.038 (-0.027 to 0.104) | 0.237 | 1.000 |
| Stunted (0/1) | 1148 | 959 | 0.061 | -0.011 (-0.041 to 0.018) | -0.011 (-0.040 to 0.018) | 0.451 | 1.000 |
| Underweight (0/1) | 1148 | 951 | 0.014 | -0.005 (-0.018 to 0.009) | -0.005 (-0.019 to 0.009) | 0.526 | 1.000 |
| Overweight (0/1) | 1148 | 946 | 0.152 | -0.016 (-0.061 to 0.028) | -0.017 (-0.061 to 0.028) | 0.467 | 1.000 |
| Any illness (0/1) | 1148 | 1148 | 0.643 | -0.082*** (-0.140 to -0.024) | -- | 0.006 | 0.036 |
| Any unintended injuries (0/1) | 1148 | 1148 | 0.092 | 0.008 (-0.028 to 0.043) | -- | 0.680 | 1.000 |
| Perinatal Mental Health<br>(Mothers) |  |  |  |  |  |  |  |
| Possible perinatal depression (0/1) | 1148 | 827 | 0.178 | -0.053** (-0.098 to -0.009) | -0.059** (-0.108 to -0.011) | 0.019 | 0.030 |
| Suicidal thoughts (0/1) | 1148 | 827 | 0.056 | -0.009 (-0.038 to 0.019) | -0.014 (-0.047 to 0.019) | 0.519 | 0.210 |
| EPDS score | 1148 | 827 | 5.212 | -0.788** (-1.391 to -0.184) | -0.897*** (-1.520 to -0.273) | 0.011 | 0.030 |
| Caregiver Mental Health<br>(Primary Caregivers) |  |  |  |  |  |  |  |
| Depression (0/1) | 1148 | 973 | 0.169 | -0.063*** (-0.108 to -0.018) | -0.061*** (-0.106 to -0.016) | 0.006 | 0.025 |
| Anxiety (0/1) | 1148 | 973 | 0.171 | -0.049** (-0.089 to -0.009) | -0.047** (-0.086 to -0.007) | 0.017 | 0.027 |
| Stress (0/1) | 1148 | 973 | 0.120 | -0.042** (-0.080 to -0.003) | -0.040** (-0.078 to -0.002) | 0.034 | 0.027 |
| DASS-21 score | 1148 | 973 | 13.598 | -2.376** (-4.555 to -0.196) | -2.302** (-4.465 to -0.139) | 0.033 | 0.027 |

Perinatal mental health for mothers was measured by the Edinburgh Postnatal Depression Scale (EPDS) and mental health for caregivers was measured by the Depression, Anxiety, and Stress Scale - 21 Items (DASS-21), with higher scores indicating greater symptom severity. All regressions included county fixed effects (randomization strata) and controlled for child-, caregiver-, and household-level covariates selected using a post-double-selection lasso procedure. Standard errors were clustered at the township level. For outcomes with incomplete follow-up data, we implemented a machine-learning lasso-based inverse probability weighting (IPW) approach and reported both intent-to-treat (ITT) estimates and IPW-adjusted estimates to assess the sensitivity of results to differential outcome completion. False discovery rate (FDR)-adjusted P-values were reported for multiple-inference correction among multiple related measures within the same domain. P-values correspond to the ITT estimates. \*\* p<0.05 \*\*\* p<0.01

By contrast, the intervention yielded significant improvements in mental health outcomes. Among mothers assessed using the Edinburgh Postnatal Depression Scale (EPDS), the prevalence of possible perinatal depression decreased by 5.3 percentage points (CI: 9.8 to 0.9; p=0.019; IPW-adjusted: 5.9), and mean EPDS scores declined by 0.788 points (CI: 1.391 to 0.184; p=0.011; IPW-adjusted: 0.897), indicating improved perinatal mental health. The EPDS is a widely validated 10-item instrument for screening perinatal depressive symptoms, with total scores ranging from 0 to 30 and a commonly used cutoff of ≥10 indicating possible perinatal depression (*31*).

Among primary caregivers assessed using the Depression, Anxiety, and Stress Scale – 21 Items (DASS-21), the intervention significantly reduced the prevalence of depressive symptoms by 6.3 percentage points (CI: 10.8 to 1.8; p=0.006; IPW-adjusted: 6.1), anxiety symptoms by 4.9 percentage points (CI: 8.9 to 0.9; p=0.017; IPW-adjusted: 4.7), and stress symptoms by 4.2 percentage points (CI: 8.0 to 0.3; p=0.034; IPW-adjusted: 4.0). Consistent with these findings, the mean DASS-21 composite scores declined by 2.376 points (CI: 4.555 to 0.196; p=0.033; IPW-adjusted: 2.302), reflecting overall improvements in caregiver mental health. The DASS-21 is a validated 21-item instrument assessing symptoms across three domains: depression, anxiety, and stress, with subscale scores ranging from 0 to 42 (*32*). Elevated symptoms were defined using standard cut-points (depression ≥10, anxiety ≥8, stress ≥15). All statistically significant effects remained significant at the 5% level after FDR adjustment for multiple hypothesis testing, and the corresponding FDR-adjusted p-values were reported.

### Effects on Secondary Infant and Young Child Feeding Outcomes

The intervention also led to significant improvements in several breastfeeding initiation indicators (Table 5). The intervention significantly increased the likelihood that children were ever breastfed by 3.2 percentage points (CI: 1.1 to 5.3; p=0.002), and that newborns received colostrum by 4.4 percentage points (CI: 0.1 to 8.6; p=0.044).

**Table 5.** Intervention effects on secondary infant and young child feeding outcomes.

|  | N<br>(eligible) | N<br>(obs) | Control<br>Mean | Overall Intervention Effect |  |  |
| --- | --- | --- | --- | --- | --- | --- |
|  |  |  |  | ITT Estimate<br>(95% CI) | P-value | FDR-adjusted<br>P-value |
| **Breastfeeding Initiation (All children)** |  |  |  |  |  |  |
| Early initiation of breastfeeding (0/1) | 1148 | 1148 | 0.189 | 0.049 (-0.001 to 0.098) | 0.052 | 0.056 |
| Newborn given colostrum (0/1) | 1148 | 1148 | 0.824 | 0.044** (0.001 to 0.086) | 0.044 | 0.056 |
| Children ever breastfed (0/1) | 1148 | 1148 | 0.937 | 0.032*** (0.011 to 0.053) | 0.002 | 0.010 |
| Exclusively breastfed first 2 days (0/1) | 1148 | 1148 | 0.100 | 0.015 (-0.025 to 0.054) | 0.470 | 0.134 |
| **Breastfeeding Practice (Infants <6 months)** |  |  |  |  |  |  |
| Mixed milk feeding (0/1) | 210 | 210 | 0.203 | -0.037 (-0.127 to 0.053) | 0.417 | -- |
| **Complementary Feeding (Children >6 months)** |  |  |  |  |  |  |
| Minimum dietary diversity (0/1) | 938 | 938 | 0.505 | 0.041 (-0.017 to 0.098) | 0.168 | 0.145 |
| Minimum meal frequency (0/1) | 938 | 938 | 0.692 | 0.031 (-0.023 to 0.085) | 0.263 | 0.188 |
| Minimum acceptable diet (0/1) | 938 | 938 | 0.381 | 0.042 (-0.013 to 0.098) | 0.136 | 0.145 |
| Consumption of egg and/or flesh food (0/1) | 938 | 938 | 0.624 | 0.059** (0.008 to 0.109) | 0.023 | 0.061 |
| Consumption of vegetable or fruits (0/1) | 938 | 938 | 0.763 | 0.054** (0.011 to 0.096) | 0.013 | 0.061 |

All regressions included county fixed effects (randomization strata) and controlled for child-, caregiver-, and household-level covariates selected using a post- double-selection lasso procedure. Standard errors were clustered at the township level. False discovery rate (FDR)-adjusted P-values were reported for multiple- inference correction among multiple related measures within the same domain. P-values correspond to the intent-to-treat (ITT) estimates. \*\* p<0.05 \*\*\* p<0.01

Improvements were also observed in several complementary feeding practices among children older than 6 months (Table 5). The intervention significantly increased the probability that children consumed eggs and/or flesh foods by 5.9 percentage points (CI: 0.8 to 10.9; p=0.023), and fruits and/or vegetables by 5.4 percentage points (CI: 1.1 to 9.6; p=0.013). All reported effects remained statistically significant at the 10% level after adjusting for multiple outcome testing within domains.

### Effects on Hygiene, Healthcare Utilization, and Prenatal Care

Table 6 presents the intervention effects on hygiene behaviors among primary caregivers, healthcare utilization among children, the number of prenatal care visits during the most recent pregnancy, maternal dietary practices among currently breastfeeding mothers, and uptake of prenatal supplements during pregnancy. No statistically significant effects were observed on these outcomes.

**Table 6.**
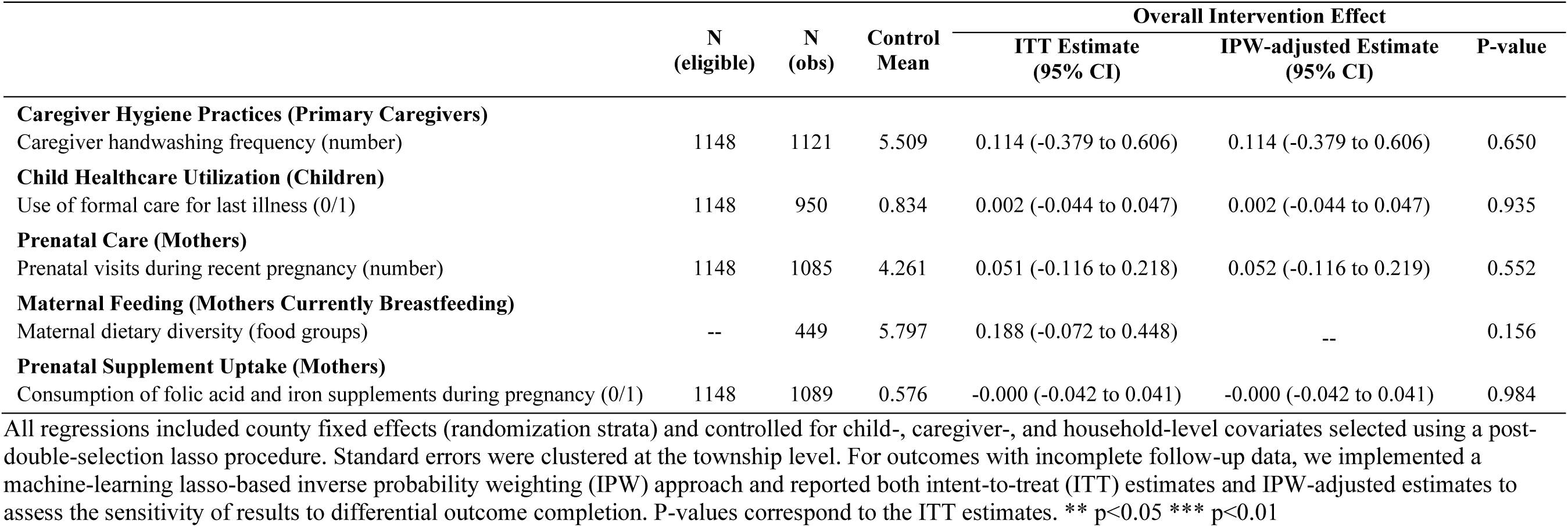
Intervention effects on hygiene, healthcare utilization, and prenatal care.

|  | N<br>(eligible) | N<br>(obs) | Control<br>Mean | Overall Intervention Effect |  | P-value |
| --- | --- | --- | --- | --- | --- | --- |
|  |  |  |  | ITT Estimate<br>(95% CI) | IPW-adjusted Estimate<br>(95% CI) |  |
| **Caregiver Hygiene Practices (Primary Caregivers)** |  |  |  |  |  |  |
| Caregiver handwashing frequency (number) | 1148 | 1121 | 5.509 | 0.114 (-0.379 to 0.606) | 0.114 (-0.379 to 0.606) | 0.650 |
| **Child Healthcare Utilization (Children)** |  |  |  |  |  |  |
| Use of formal care for last illness (0/1) | 1148 | 950 | 0.834 | 0.002 (-0.044 to 0.047) | 0.002 (-0.044 to 0.047) | 0.935 |
| **Prenatal Care (Mothers)** |  |  |  |  |  |  |
| Prenatal visits during recent pregnancy (number) | 1148 | 1085 | 4.261 | 0.051 (-0.116 to 0.218) | 0.052 (-0.116 to 0.219) | 0.552 |
| **Maternal Feeding (Mothers Currently Breastfeeding)** |  |  |  |  |  |  |
| Maternal dietary diversity (food groups) | -- | 449 | 5.797 | 0.188 (-0.072 to 0.448) | -- | 0.156 |
| **Prenatal Supplement Uptake (Mothers)** |  |  |  |  |  |  |
| Consumption of folic acid and iron supplements during pregnancy (0/1) | 1148 | 1089 | 0.576 | -0.000 (-0.042 to 0.041) | -0.000 (-0.042 to 0.041) | 0.984 |

All regressions included county fixed effects (randomization strata) and controlled for child-, caregiver-, and household-level covariates selected using a post- double-selection lasso procedure. Standard errors were clustered at the township level. For outcomes with incomplete follow-up data, we implemented a machine-learning lasso-based inverse probability weighting (IPW) approach and reported both intent-to-treat (ITT) estimates and IPW-adjusted estimates to assess the sensitivity of results to differential outcome completion. P-values correspond to the ITT estimates. \*\* p<0.05 \*\*\* p<0.01

### Effects on Intermediate Outcomes

Table 7 presents the intervention effects of the intervention on potential intermediate outcomes, including caregiving knowledge and perceived social support among primary caregivers. Caregiving knowledge was assessed using a broad set of questions covering breastfeeding, complementary feeding, child illness management, and caregiver mental health self-management. Responses were aggregated into indices, including an overall caregiving knowledge index and a mental health self-management knowledge index. Perceived social support was measured using the Multidimensional Scale of Perceived Social Support, with mean subscale scores ranging from 1 to 7 across three subscales (family, friends, and significant others), where higher values indicate greater perceived support (*33*).

**Table 7.**
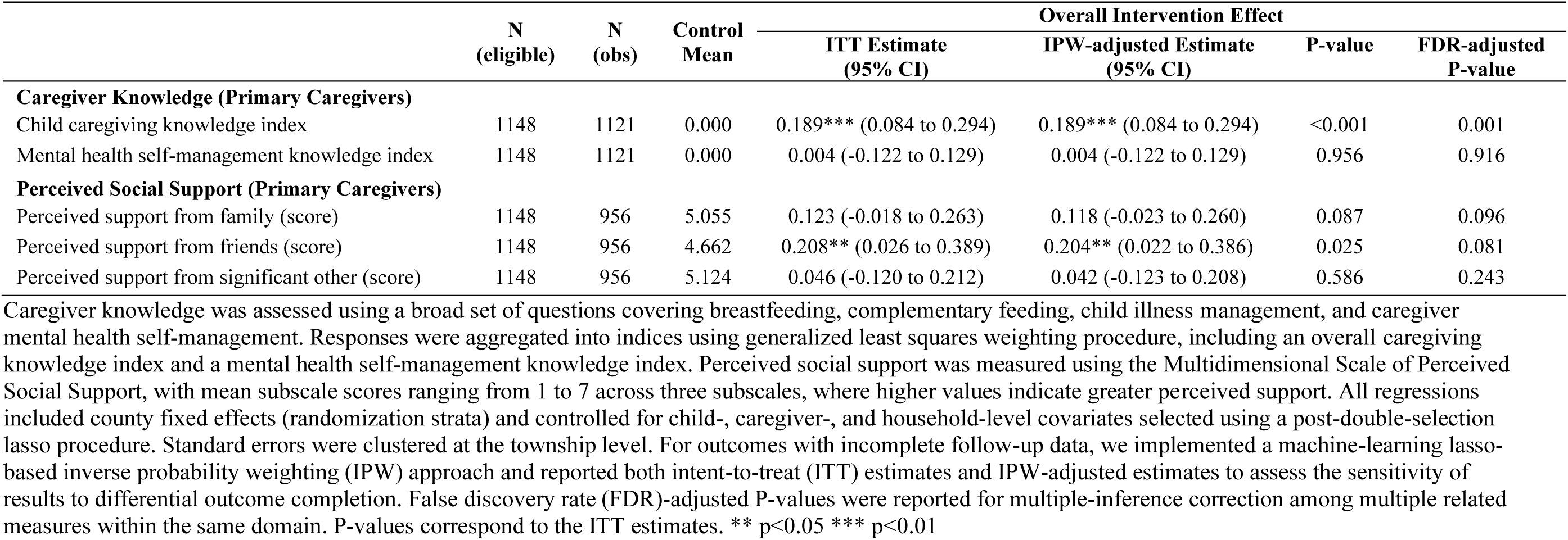
Intervention effects on intermediate outcomes.

|  | N<br>(eligible) | N<br>(obs) | Control<br>Mean | Overall Intervention Effect |  |  |  |
| --- | --- | --- | --- | --- | --- | --- | --- |
|  |  |  |  | ITT Estimate<br>(95% CI) | IPW-adjusted Estimate<br>(95% CI) | P-value | FDR-adjusted<br>P-value |
| **Caregiver Knowledge (Primary Caregivers)** |  |  |  |  |  |  |  |
| Child caregiving knowledge index | 1148 | 1121 | 0.000 | 0.189*** (0.084 to 0.294) | 0.189*** (0.084 to 0.294) | <0.001 | 0.001 |
| Mental health self-management knowledge index | 1148 | 1121 | 0.000 | 0.004 (-0.122 to 0.129) | 0.004 (-0.122 to 0.129) | 0.956 | 0.916 |
| **Perceived Social Support (Primary Caregivers)** |  |  |  |  |  |  |  |
| Perceived support from family (score) | 1148 | 956 | 5.055 | 0.123 (-0.018 to 0.263) | 0.118 (-0.023 to 0.260) | 0.087 | 0.096 |
| Perceived support from friends (score) | 1148 | 956 | 4.662 | 0.208** (0.026 to 0.389) | 0.204** (0.022 to 0.386) | 0.025 | 0.081 |
| Perceived support from significant other (score) | 1148 | 956 | 5.124 | 0.046 (-0.120 to 0.212) | 0.042 (-0.123 to 0.208) | 0.586 | 0.243 |

Caregiver knowledge was assessed using a broad set of questions covering breastfeeding, complementary feeding, child illness management, and caregiver mental health self-management. Responses were aggregated into indices using generalized least squares weighting procedure, including an overall caregiving knowledge index and a mental health self-management knowledge index. Perceived social support was measured using the Multidimensional Scale of Perceived Social Support, with mean subscale scores ranging from 1 to 7 across three subscales, where higher values indicate greater perceived support. All regressions included county fixed effects (randomization strata) and controlled for child-, caregiver-, and household-level covariates selected using a post-double-selection lasso procedure. Standard errors were clustered at the township level. For outcomes with incomplete follow-up data, we implemented a machine-learning lasso- based inverse probability weighting (IPW) approach and reported both intent-to-treat (ITT) estimates and IPW-adjusted estimates to assess the sensitivity of results to differential outcome completion. False discovery rate (FDR)-adjusted P-values were reported for multiple-inference correction among multiple related measures within the same domain. P-values correspond to the ITT estimates. \*\* p<0.05 \*\*\* p<0.01

The intervention significantly improved the overall caregiving knowledge index among primary caregivers, with an increase of 0.189 standard deviations (SD) (CI: 0.084 to 0.294; p<0.001; IPW-adjusted: 0.189), which may help contextualize the observed improvements in several caregiving behaviors. With respect to caregiver mental health, we found no statistically significant effects on the knowledge index related to mental health self-management. In contrast, the intervention significantly increased perceived social support, particularly support reported from friends, by 0.208 points (CI: 0.026 to 0.389; p=0.025; IPW-adjusted: 0.204). These findings suggest that improvements in caregiver mental health may have been primarily associated with enhanced perceived social support rather than increased mental health self-management knowledge. Both effects remained statistically significant at the 10% level after adjusting for multiple outcome testing within their respective domains.

### Effects Based on Domain-level Index Outcomes

Figure 4 plots the intervention effects on domain-level summary indices, with higher values indicating better outcomes. Because the *Healthy Future* program is a comprehensive, multi-component intervention, these domain-level summary tests were added post-hoc to allow a more comprehensive assessment of program effects across domains, complementing the analyses based on individual outcome measures. The same index construction procedure was applied consistently across all domains for comparison of intervention effect estimates.

**Figure 4.**
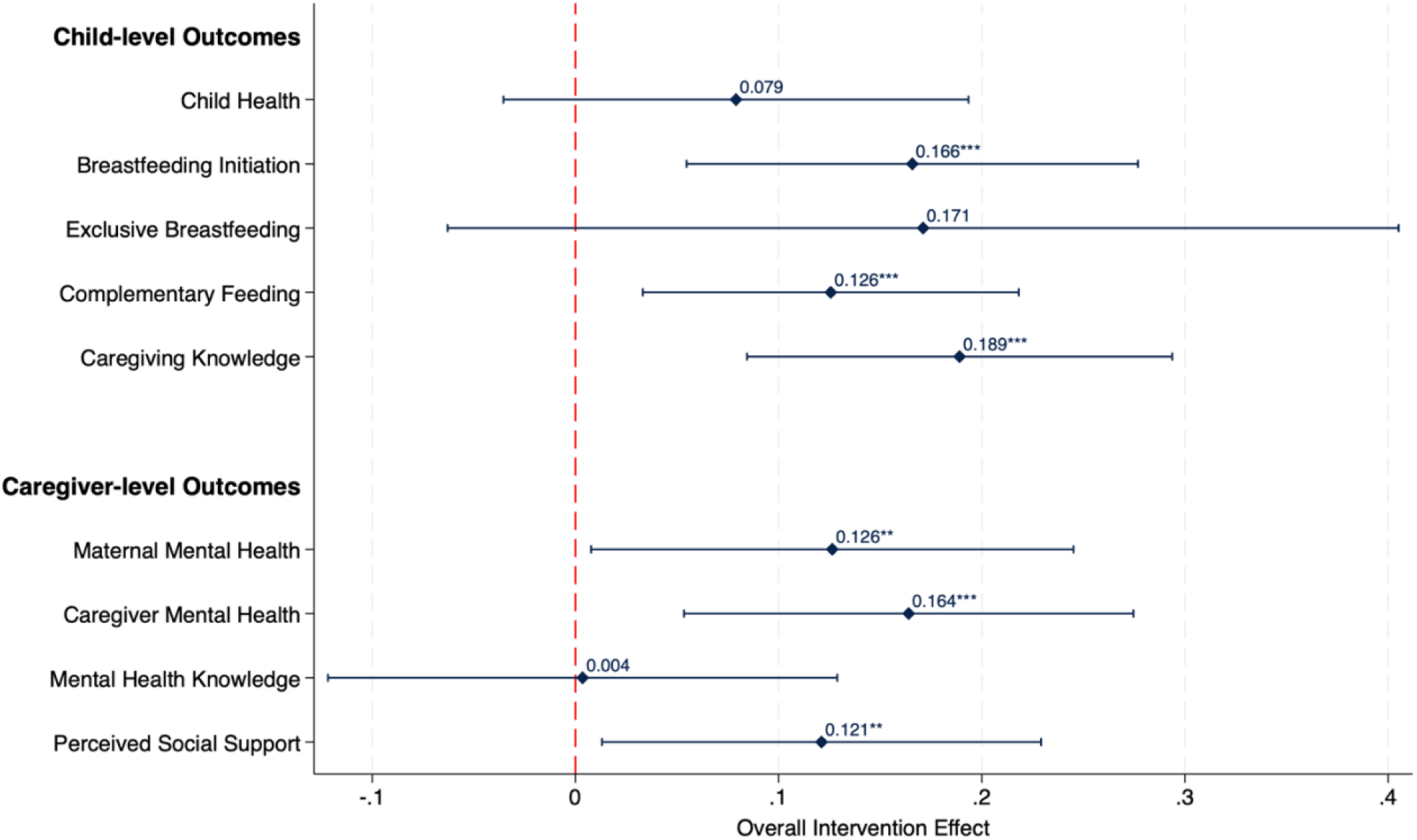
Intervention effects on domain-level summary indices. This figure presents standardized treatment effects on domain-level summary indices constructed from aggregating related primary and secondary outcomes within each outcome domain. Points represent estimated average treatment effects and bars indicate 95% confidence intervals. Higher values indicate better outcomes. All regressions included county fixed effects (randomization strata) and controlled for child-, caregiver-, and household-level covariates selected using a post-double-selection lasso procedure. Standard errors were clustered at the township level. ** p<0.05 *** p<0.01

By combining related primary and secondary outcomes into domain-level summary indices, we found no statistically significant improvement in the overall child health index (0.079 SD; CI: −0.035 to 0.193; p=0.176), while observing significant improvements in breastfeeding initiation index (0.166 SD; CI: 0.055 to 0.277; p=0.003) and complementary feeding index (0.126 SD; CI: 0.033 to 0.218; p=0.008). The absence of statistically significant effects on exclusive breastfeeding index (0.171 SD; −0.063 to 0.405; p=0.152) largely reflects limited statistical power among infants younger than 6 months at follow-up, resulting in a wider confidence interval. These behavioral improvements were accompanied by increased caregiver knowledge index (0.189 SD; CI: 0.084 to 0.294; p<0.001).

In the mental health domain, domain-based analyses indicate significant improvements in mental health indices among mothers (0.126 SD; CI: 0.008 to 0.245; p=0.037) and among primary caregivers (0.164 SD; CI: 0.053 to 0.275; p=0.004). Consistent with findings from the intermediate outcomes analysis, these improvements align more closely with increases in perceived social support index (0.121 SD; CI: 0.013 to 0.229; p=0.028) than with changes in mental health self-management knowledge index (0.004 SD; CI: −0.122 to 0.129; p=0.956). This pattern may suggest that regular CHW engagement through structured home visits have functioned as an important source of social support, alongside the provision of informational content.

Overall, these domain-level summary indices suggest that the intervention’s integrated curriculum design and delivery model may have contributed to coherent improvements across behavioral and mental health outcomes, even when effects on individual outcome measures varied.

### Results from the Compliance-based TOT Analysis

Results from the compliance-adjusted TOT analysis are reported in Appendix Section S4. These estimates were obtained using an IV approach, with assignment to an intervention township serving as an instrument for the cumulative number of completed home visits before the follow-up survey. We report these results as sensitivity analyses to complement the ITT estimates presented in the main text. Overall, the compliance-based TOT estimates were generally consistent with the ITT findings and led to similar substantive conclusions, although these two approaches differ in interpretation: ITT estimates capture the overall effects of offering the *Healthy Future* program, whereas TOT estimates capture the effects of intervention exposure among participating families.

## Discussion

In this study, we present how we integrated a digitally-enabled system to operationalize comprehensive and flexible maternal and child intervention designs in low-resource settings, and whether such integration can translate into meaningful improvements across multidimensional maternal and child health outcomes. To address the holistic and evolving health needs of mothers and children during the critical first 1,000 days of life, we designed and evaluated a comprehensive yet flexible program, the *Healthy Future* program, featuring a stage-based maternal and child curriculum, integrated with digital technology to address logistical challenges and enable developmentally appropriate support, and delivered through a well-established CHW-led home-visiting model in low-resource settings. Together, this study demonstrates our delicate effort to move beyond stand-alone interventions toward a multidimensional delivery model that is comprehensive, responsive to families’ evolving needs, and feasible for large-scale implementation in resource-constrained settings. We evaluated the effect of the *Healthy Future* program through a cluster-randomized controlled trial across 119 rural townships in four counties in Sichuan Province, China, previously designated as national poverty-stricken areas.

The intervention did not significantly improve two of the three primary outcomes, hemoglobin levels and exclusive breastfeeding, although dietary diversity improved modestly. The lack of a significant effect on exclusive breastfeeding may have been partly attributable to the consolidation of two follow-up surveys caused by COVID-19 disruptions, which reduced the number of infants younger than six months available for analysis and consequently reduced statistical power. By contrast, the absence of an effect on hemoglobin concentration may reflect the difficulty of influencing anemia-related outcomes within the relatively short timeframe of the intervention, which only lasted 12 months.

However, the three primary outcomes were concentrated in the domain of child nutrition, whereas the *Healthy Future* program was designed as a comprehensive intervention targeting multiple dimensions of maternal and child health. Consistent with this broad scope, we also examined a broad range of secondary outcomes, including maternal and caregiver mental health, maternal nutrition, hygiene practices, caregiving knowledge, and perceived social support. Given the intervention’s multi-component design, we examined both individual outcomes and domain-level summary indices constructed from the pre-specified set of primary and secondary outcomes. Domain-level indices provide a high-level summary of intervention effects across domains.

Viewed together, our analyses suggest that improvements observed across multiple related indicators may have translated into coherent patterns of gains within several targeted domains. Specifically, we observed significant improvements in breastfeeding initiation and complementary feeding practices, as well as increased caregiving knowledge. The program also yielded substantial improvements in maternal and caregiver mental health, including reductions in symptoms of depression, anxiety, and stress, alongside increased perceived social support. This pattern suggests that regular CHW engagement through structured home visits may have contributed to the observed mental health benefits by providing consistent interpersonal contact, emotional reassurance, and opportunities for social connection, alongside the provision of informational content. In contrast, no significant effects were observed on child health outcomes or exclusive breastfeeding practices.

Overall, the observed pattern of findings is broadly consistent with the intervention’s theory of change. The strongest effects were observed in the domains most proximal to the mechanisms directly targeted by the intervention, including caregiving knowledge, perceived social support, complementary feeding practices, or caregiver mental health. By contrast, effects on more biological outcomes were limited during the study period, suggesting that changes in downstream child health outcomes may require longer exposure and follow-up to emerge.

This study represents our broad efforts and dedications to design comprehensive and flexible intervention to address the multidimensional and evolving challenges faced by mothers and children during the first 1,000 days of life, a difficult but important problem that often cannot be adequately addressed through narrowly focused interventions. First, the curriculum we designed was extensive yet flexible, crafted to meet the diverse needs of mothers and children during critical developmental periods. It was stage-based, tailored to the evolving needs of families, and adaptable to varying times of entry into the program. This structured yet dynamic approach ensured that the guidance and support provided were both relevant and immediately applicable. Second, our intervention capitalized on the globally accepted practice of home visits by CHWs, enhancing the adaptability of the program to diverse settings. This delivery method is well-established and widely recognized for its effectiveness in engaging families in difficult-to-reach regions in various settings (*47–49*). Third, the integration of a tablet-based app for CHWs was a pivotal component of our strategy to manage the complexities involved in delivering comprehensive and flexible interventions. This app was meticulously aligned with the curriculum to automate assigning highly scripted content with detailed instructions, significantly reducing logistical challenges for CHWs, and ensuring consistent delivery of materials while also enabling CHWs to identify and fill gaps in family knowledge. Moreover, the app captured reliable process information, enabling real-time tracking and active supervisory monitoring of the frequency and length of CHW home visits.

We conducted an initial search of published literature in developing our study protocol and performed an updated formal search on PubMed in preparation for this manuscript. Search terms included “intervention,” “community health worker,” “child,” “mother,” “1000 days,” “comprehensive,” “integrated,” “stage-based,” “adaptive,” and “tailored.” Inclusion criteria were studies conducted in developing countries or low-resource settings, involving interventions delivered by CHWs targeting children and mothers from pregnancy to infant age two years, designed as comprehensive/integrated and stage-based/adaptive, with or without the assistance of digital technology. We identified no published literature with similar intervention designs, highlighting the innovative nature of our approach and its potential to fill a critical gap in maternal and child health research globally. While our literature search identified a few integrated programs aimed at addressing the diverse challenges that mothers and children face (*48*, *50–53*), most focused on testing the feasibility of combining multiple health initiatives, and few were designed to be adaptive like ours. Furthermore, these programs often highlighted the significant logistical challenges CHWs encounter when delivering interventions, emphasizing the need for innovative solutions to promote scalability. Digital technology has been increasingly used in maternal and child programs (*24–26*, *54*, *55*). However, most digital innovations relied on “push” technology, digital reminders, or audio/video messages (*24*). In contrast, our program took a distinctly different approach by meticulously designing an app specifically crafted to integrate with the intervention design to support CHWs in their fieldwork. Rather than serving solely as a communication channel, the app was coherently designed to reduce the workload and logistical complexity associated with timely content delivery, thereby improving the consistency and fidelity of intervention implementation. This strategic use of digital technology sets our approach apart from less integrated digital solutions, highlighting its uniqueness and potential for broader applications.

A key limitation of this study is that the trial evaluation was not ideal. COVID-19 disruptions during the study period substantially affected the original evaluation plan. Repeated lockdowns and quarantine requirements necessitated the consolidation of the two planned follow-up surveys into one extended follow-up survey, resulting in several deviations from the original protocol and creating challenges for evaluating some age-specific outcomes, particularly exclusive breastfeeding, one of the primary outcomes. The disruption also contributed to differential completion rates for some outcome measures. Although we implemented inverse probability weighting and other sensitivity analyses to assess the robustness of the findings, these approaches rely on assumptions that may not fully account for all sources of bias. Consequently, the results should be interpreted while considering these implementation and data collection challenges. However, we note that COVID-19 restrictions primarily affected data collection rather than program implementation, as large enumerator teams were subject to travel restrictions, while CHWs’ engagement with families was flexible and could be easily adjusted with a flexible program design.

A second limitation relates to the evaluation of a comprehensive, multi-component intervention targeting multiple dimensions of maternal and child health challenges. Such interventions may generate modest effects across a broad range of outcomes rather than large effects on any single indicator (*11*), making interpretation challenging when numerous related outcomes are examined. To facilitate interpretation, we added domain-level summary indices post-hoc that aggregated related primary and secondary outcomes within each domain. This index construction was not specified in the original protocol and should therefore be interpreted cautiously. Nevertheless, this approach has been increasingly used in the evaluation of complex interventions to provide a broader perspective on intervention impacts (*44–46*). Moreover, as advocacy grows for integrated interventions that address the holistic needs of at-risk families (*6*, *9*), there is a clear need for further research into the effects of such interventions across a wide range of outcomes, recognizing that limited effects may emerge on single outcomes.

This study also has several other limitations that highlight areas for future research. First, our program focused on the period from pregnancy up to child age 18 months, primarily emphasizing health and nutrition for mothers and children. It did not incorporate significant opportunities for early learning during early childhood, another critical area for supporting children’s development (*7*). Future research could explore innovative approaches to delivering and scaling interventions that integrate nutrition and early stimulation, such as how to optimize these inputs to promote developmental outcomes for children of all ages (*11*). Second, our adaptive design was primarily centered on developmental stages, while allowing CHWs to manually supplement the content with additional customized materials to meet specific family needs. Future research could explore more highly personalized interventions that consider both the developmental stages and the circumstances of individual families, enhancing the specificity and effectiveness of the interventions. Lastly, this paper focused primarily on overall program effects, but our ongoing work aims to explore more nuanced effects, such as heterogeneous effects across different characteristics. This continued research will further elucidate how integrated and flexible health interventions can be optimized for maximum impact across a diverse population.

Overall, our study supports the potential to design comprehensive and flexible interventions while mitigating implementation complexities through the integration of digital health technology. Most importantly, our flexible, community-based model represents an effort to complement national maternal and child health initiatives, which often focus on improving population-level outcomes while overlooking the complex and nuanced needs of families, particularly those living in low-resource communities. With growing advocacy for comprehensive approaches to holistically address the diverse and evolving health needs of mothers and children during critical developmental periods, our study aims to address a central question: how such interventions can be designed, implemented, and scaled in ways that ensure timely support for families without overstretching frontline health workers. Such research is essential for advancing the understanding of how to enhance the reach, relevance, and sustainability of health interventions that improve outcomes for mothers and children globally, while ensuring that underserved families receive timely and appropriate support when they need it most.

## Data Availability

All data produced in the present study are available upon reasonable request to the authors.

## Acknowledgments

We thank the participants of the *Healthy Future* program, the local partners, the enumerator team, and the community health workers for their valuable contributions and dedication to this study.

## Author contributions

Conceptualization: SS, SR, GLD, AMW, AM, HZ

Methodology: SS, AMW, GLD, YC

Funding acquisition: SS, SR, AM, YG, GLD, HZ

Project administration: AM, YG, YW, YC

Investigation: YW, HZ

Supervision: SS

Formal analysis: YC

Writing—original draft: YC

Writing—review & editing: SS, GLD, AMW, SB, HWZ

## Data and materials availability

All data and replication codes for this paper will be available in a public repository before publication.

## Supplementary Materials

### S1. Healthy Future Program Design, Enrollment, and Completion

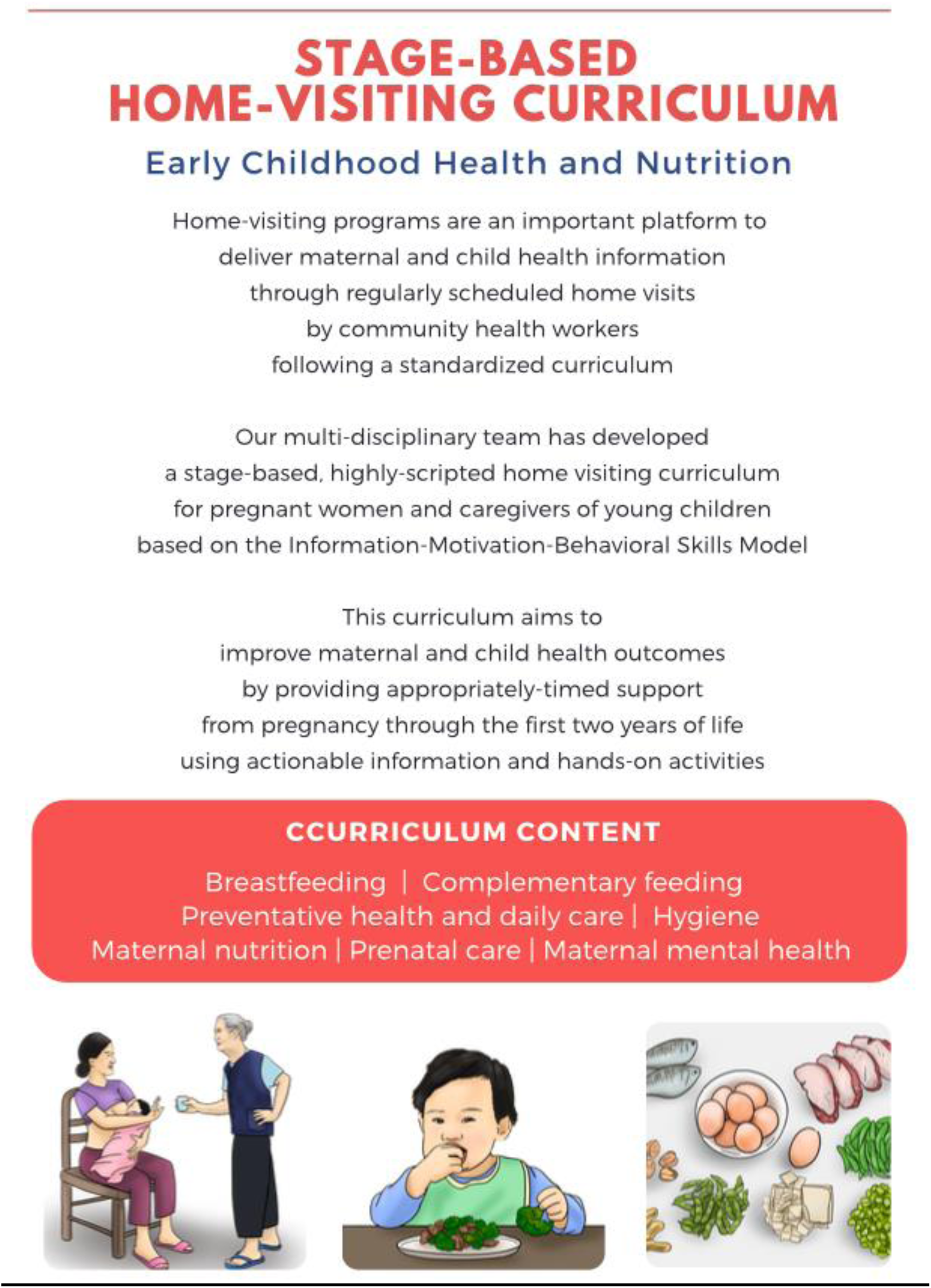

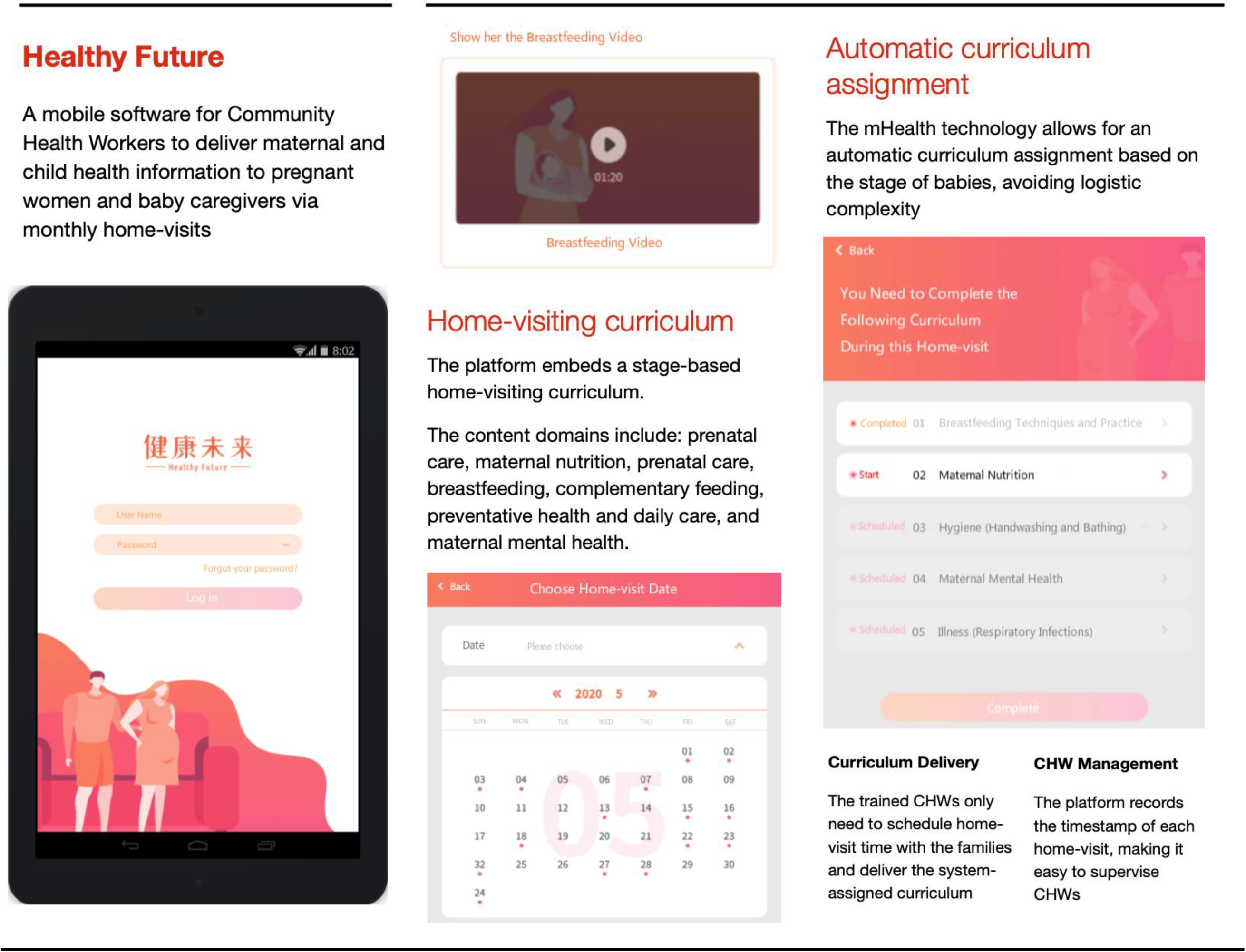

#### Healthy Future Curriculum: Table of Contents

**Pregnancy 4-6 Months**

Session H4

H4.1: Maternal Nutrition (Food groups, iron and anemia) 5

H4.2: Benefits and Importance of Breastfeeding (Intro & concerns) 10

H4.3: Caregiver Mental Health (Three Pillars Intro) 14

H4.4: Uptake of Government Services (Maternal Health Check-ups) 20

H4 Knowledge/Attitude Check 21

Session H5 23

H5.1: Maternal nutrition (Micronutrients and exercise) 23

H5.2: Caregiver Mental Health (Healthy Thinking) 27

H5 Knowledge/Attitude Check 31

Session H6 32

H6.1: Maternal Nutrition (Water and dangerous substances) 32

H6.2: Benefits and Importance of Breastfeeding (Signs of hunger, breastfeeding when ill) 35

H6.3: Caregiver Mental Health (Slow Breathing Exercise) 37

H6 Knowledge/Attitude Check 39

**Pregnancy 7-9 Months**

Session H7 40

H7.1: Breastfeeding Techniques and Practice 40

H7.2: Maternal Nutrition (Review food diversity and iron) 46

H7.3: Caregiver Mental Health (Social Support) 49

H7 Knowledge/Attitude Check 51

Session H8 52

H8.1: Breastfeeding (Breastfeeding Support) 52

H8.2: Maternal Nutrition (Review micronutrients, blood pressure, and diabetes) 54

H8.3: Illness Prevention 56

H8 Knowledge/Attitude Check 58

Session H9 59

H9.1: Breastfeeding (Frequency and length of time) 59

H9.2: Maternal Nutrition (Review water and dangerous substances) 62

H9.3: Uptake of Government Service (Baby Well-visits) 64

H9 Knowledge/Attitude Check 65

**INFANCY 1 Month**

Infancy 0-6 months Special Module: Formula Feeding 66

Session B1-1 70

B1-1.1: Breastfeeding Techniques and Practice (Troubleshooting) 70

B1-1.2: Maternal Nutrition (During breastfeeding) 76

B1-1.3: Hygiene (Handwashing, bathing and cord care) 79

B1-1.4: Caregiver Mental Health (Understanding Mental Health) 81

B1-1 Knowledge/Attitude Check 82

Session B1-2 83

B1-2.1: Breastfeeding Check-in (Pain Troubleshooting) 83

B1-2.2: Maternal Nutrition (Review 1 if breastfeeding) 88

B1-2.3: Hygiene (Infant pee and poop) 89

B1-2.4: Caregiver Mental Health (Self-care) 91

B1-2 Knowledge/Attitude Check 95

Session B1-3 96

B1-3.1: Breastfeeding Check-in (Breastfeeding Support) 96

B1-3.2: Maternal Nutrition (Review 2 if breastfeeding) 101

B1-3.3: Hygiene (Cleanliness tips) 102

B1-3.4: Caregiver Mental Health (Baby Bonding) 104

B1-3 Knowledge/Attitude Check 107

**INFANCY 2-3 Months**

Session B2 108

B2.1: Breastfeeding Check In; Breastfeeding Support and Troubleshooting 108

B2.2: Maternal Nutrition 117

B2.3: Caregiver Mental Health (Baby Bonding Activity) 118

B2 Knowledge/Attitude Check 123

Session B3 124

B3.1: Illness (Respiratory Infections) 124

B3.2: Breastfeeding Check In (Exclusive breastfeeding) 128

B3.3: Maternal Nutrition 133

B3.4: Caregiver Mental Health (Building Confidence – newborn) 134

B3 Knowledge/Attitude Check 135

**INFANCY 4-6 Months**

Session B4 136

B4.1: Transition to Solids (Energy needs) 136

B4.2: Breastfeeding Check-in 139

B4.3: Hygiene and Illness (Constipation & Diarrhea) 146

B4.4: Caregiver Mental Health (Relaxation Techniques) 150

B4 Knowledge/Attitude Check 152

Session B5 153

B5.1: Transition to Solids (Developmental readiness) 153

B5.2: Breastfeeding Check-in 155

B5.3: Caregiver Mental Health (Mood Chart) 161

B5.4: Uptake of Government Service (YYB)163

B5 Knowledge/Attitude Check 164

Session B6 165

B6.1: Transition to Solids (Overview of what, when & how) 165

B6.2: Breastfeeding Check-in 171

B6.3: Hygiene (Review & Teeth brushing) 175

B6 Knowledge/Attitude Check 177

**INFANCY 7-9 Months**

Infancy 7-12 months Special Module: Formula Feeding 178

Session B7 179

B7.1: Transition to Solids (Snacks & meal preparation tips) 179

B7.2: Breastfeeding Check-in 182

B7 Knowledge/Attitude Check 183

Session B8 184

B8.1: Responsive Feeding 184

B8.2: Breastfeeding Check-in 187

B8.3: Uptake of Government Service – YYB Check in 188

B8 Knowledge/Attitude Check 189

Session B9 190

B9.1: Respiratory Infections Check-In 190

B9.2: Transition to Solids (Self-feeding) 192

B9.3: Breastfeeding check-in 195

B9 Knowledge/Attitude Check 196

**INFANCY 10-12 Months**

Session B10 197

B10.1: Injury (Prevention – Choking) 197

B10.2: Transition to Solids (Refresher and new textures) 199

B10.3: Breastfeeding Check-in 201

B10.4: Caregiver Mental Health (Draw Your Feelings) 202

B10.5: Uptake of Government Service – YYB Check in 203

B10 Knowledge/Attitude Check 204

Session B11 205

B11.1: Injury (Prevention - Falls & burns) 205

B11.2: Breastfeeding Check-in 207

B11.3: Caregiver Mental Health (Check-in) 208

B11 Knowledge/Attitude Check 209

Session B12 210

B12.1: Transition to Solids (Feeding after 12 months) 210

B12.2: Breastfeeding Check-in 212

B12.3: Caregiver Mental Health (Building Confidence - toddler) 213

B12 Knowledge/Attitude Check 214

**INFANCY 13-15 Months**

Session B13 215

B13.1: Injury (Prevention – Poisons) 215

B13 Knowledge/Attitude Check 217

Session B14 218

B14.1: Healthy Local Foods (Refresher – food diversity) 218

B14.2: Injury (Care-seeking: Falls, cuts & burns) 220

B14.3: Uptake of Government Service – YYB Check in 222

B14 Knowledge/Attitude Check 223

Session B15 224

B15.1: Respiratory Infections (Check-in) 224

**INFANCY 16-18 Months**

Session B16 226

B16.1: Healthy Local Foods (Refresher – meal preparation) 226

B16.2: Injury (Prevention – Review) 228

Session B17 231

B17.1: Injury (Care-seeking – Review) 231

Session B18 234

B18.1: Caregiver Mental Health (Slow Breathing Exercise) 234

**Table A.**
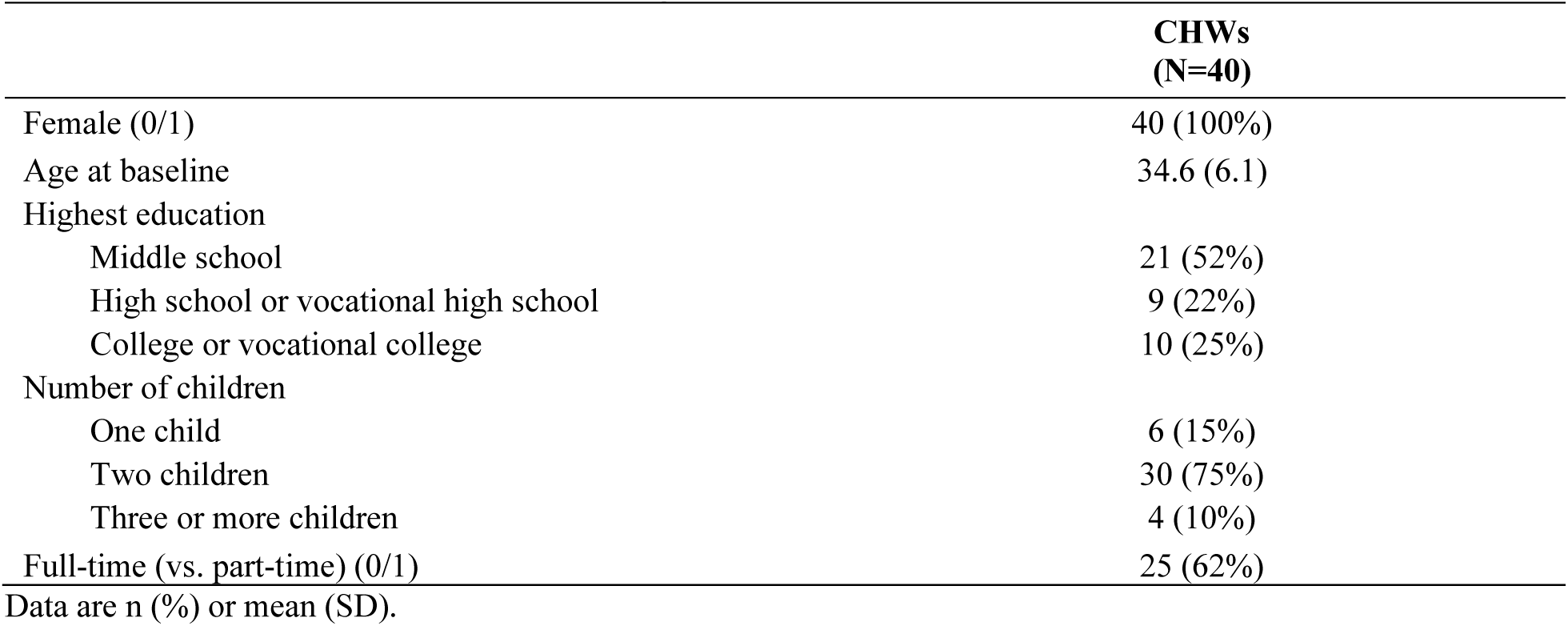
Characteristics of Community Health Workers.

|  | **CHWs**<br>**(N=40)** |
| --- | --- |
| Female (0/1) | 40 (100%) |
| Age at baseline | 34.6 (6.1) |
| Highest education |  |
| Middle school | 21 (52%) |
| High school or vocational high school | 9 (22%) |
| College or vocational college | 10 (25%) |
| Number of children |  |
| One child | 6 (15%) |
| Two children | 30 (75%) |
| Three or more children | 4 (10%) |
| Full-time (vs. part-time) (0/1) | 25 (62%) |

Data are n (%) or mean (SD).

**Figure A.**
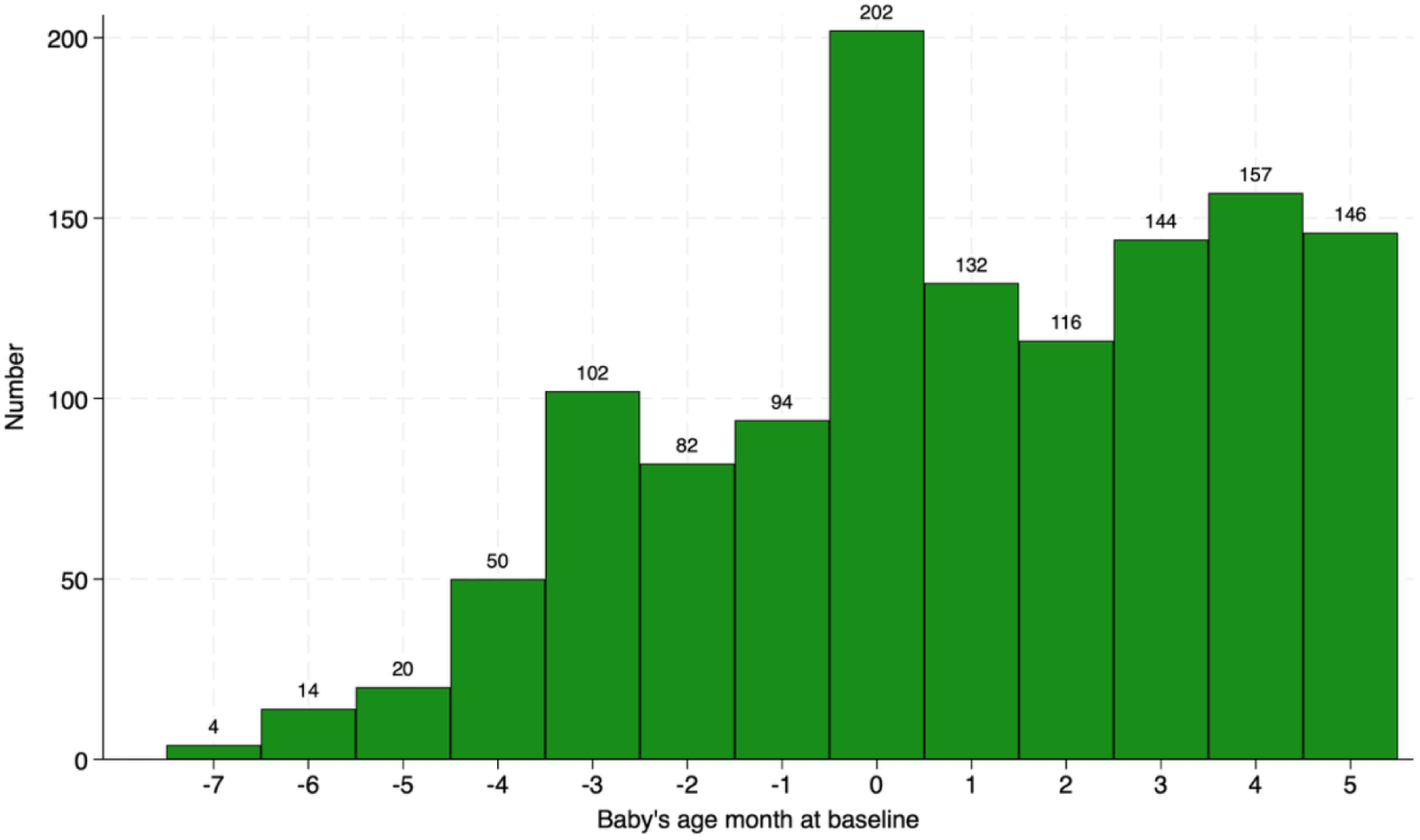
Distribution of child age at baseline.

**Figure B.**
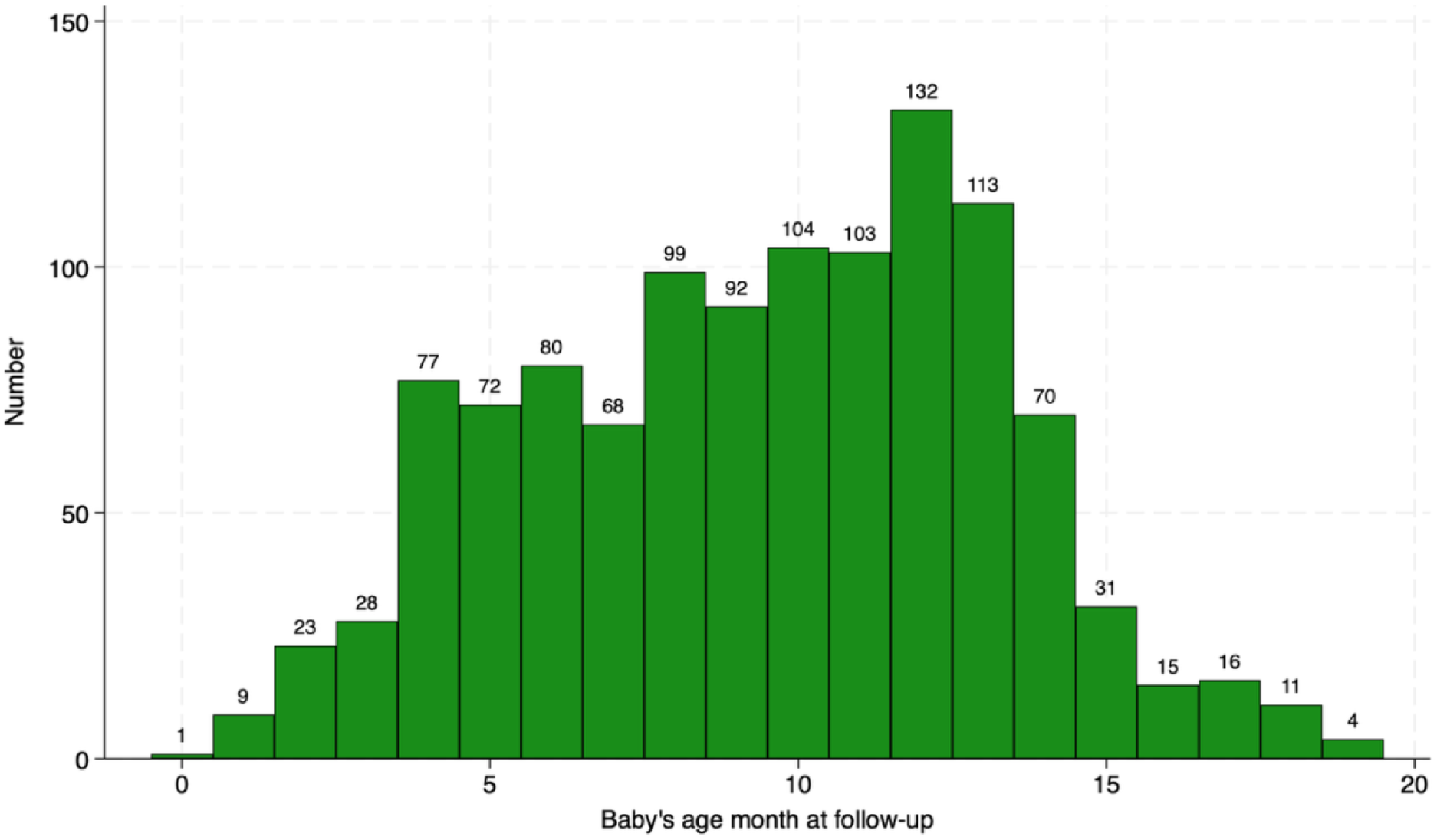
Distribution of child age at follow-up.

**Figure C.**
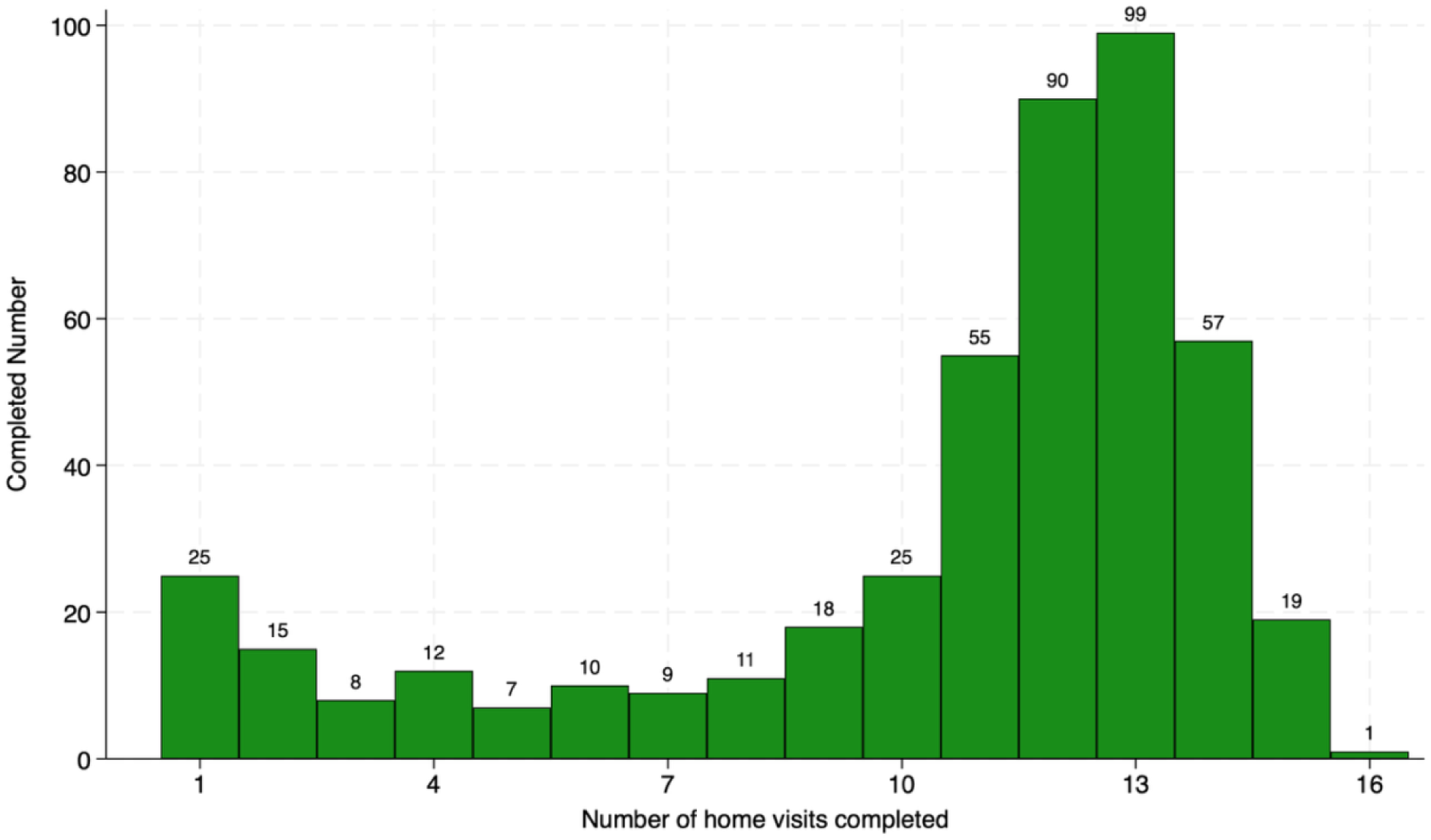
Completion of home visits by participating families. This figure plots the distribution of completed home visits among participating families over the 12-month intervention period.

### S2. Deviation from the Protocol and Study Outcome Measures

#### 1. Changes to Follow-up Survey Plan

We originally planned two follow-up surveys, as outlined in the study protocol: one in March 2022 and one in August 2022. However, the survey team experienced repeated COVID-19 lockdowns and quarantines which substantially disrupted data collection, while the intervention itself continued operating in real time as children continued aging through critical developmental stages. Under these circumstances, the study team had to make difficult decisions to preserve follow-up and intervention continuity as much as possible. Therefore, we modified the plan to prioritize achieving a high follow-up rate for one single extended follow-up survey administered in person and by telephone in two phases: March to May 2022 and August to October 2022.

Notably, COVID-19 field restrictions primarily affected data collection rather than program implementation. Data collection relied on large enumerator teams that were subject to travel restrictions and lockdowns, leading to delays and increased costs. In contrast, CHWs’ engagement with families was more flexible and could be readily adjusted. Missed visits did not disrupt subsequent intervention delivery, as the stage-based, interval-structured program design allowed content to realign with families’ current developmental stages.

#### 2. Combination of two intervention arms

As specified in the study protocol, the intervention group consisted of two delivery strategies: a standard intervention strategy primarily targeting primary caregivers in 20 intervention townships and a family-encouragement strategy encouraging secondary caregiver participation in the other 20 intervention townships. This paper combines these two intervention arms and focuses on describing the development of the *Healthy Future* program and evaluating its overall effectiveness relative to the control group. The comparative analysis of these two delivery strategies is reported separately.

We note that the only difference between the two strategies was the encouragement of secondary caregiver participation. In the standard intervention arm, home visits were scheduled primarily with pregnant women or primary caregivers of young children (typically mothers), although secondary caregivers were welcome to participate voluntarily. In the family-encouragement arm, CHWs actively encouraged the participation of secondary caregivers and attempted to schedule visits with both primary and secondary caregivers whenever possible. However, participation by secondary caregivers remained voluntary, and home visits proceeded when secondary caregivers were unavailable or unwilling to participate.

As shown in Table A, the encouragement strategy substantially increased secondary caregiver participation (45.0% versus 21.3%) but had little effect on intervention uptake or primary caregiver participation (96.7% versus 95.5%). Thus, the principal distinction between the two intervention arms was the intensity of secondary caregiver engagement rather than differences in the content or delivery of the *Healthy Future* program. Overall, the two intervention arms shared the same curriculum, delivery platform, implementation procedures, and home-visiting schedule.

#### 3. Changes to Study Outcomes

In the study protocol, we specified a broad set of secondary outcomes reflecting the integrated content domains covered by the *Healthy Future* curriculum. Table B in Appendix Section S2 outlines the full list of outcome measures and summarizes deviations from the original study protocol. Two main changes were made to the originally specified secondary outcomes.

First, several infant and young child feeding indicators were updated. At the time of trial registration and protocol construction, the outcome list was based on the 2010 WHO *Indicators for Assessing Infant and Young Child Feeding Practices* guide. However, by the time data collection began, the updated 2022 WHO guidance had been released, which removed some previous indicators and introduced new ones. Accordingly, we prioritized the use of the indicators recommended in the 2022 WHO guide where applicable in the analysis.

Second, we removed a subset of intermediate outcomes for age-specific outcomes, such as measures of breastfeeding attitudes and self-efficacy. These indicators were originally included to assess potential mediators of breastfeeding behavior. However, due to the consolidation of the two planned follow-up surveys into a single follow-up, necessitated by COVID-19 disruptions, sample sizes for key subgroups were substantially reduced. For instance, only 210 infants in the follow-up sample were aged 0–6 months for assessing many breastfeeding outcomes, and the number of observations with complete data for the corresponding intermediate outcomes was even smaller. Given limited statistical power to evaluate intervention effects on these measures, they were excluded from the final analysis.

#### 4. Addition of Domain-level Summary Indices

To facilitate a high-level interpretation of the program’s overall effects, we added analyses based on domain-level summary indices, which were not specified in the published protocol. The indices were constructed by aggregating related primary and secondary outcome measures within each domain into a single summary measure using the generalized least squares weighting procedure. Table B in Appendix S3 provides the list of primary and secondary measures used to construct the domain-level summary indices.

The main motivation for including these indices is that they provide a coherent, high-level interpretation of the program’s overall impacts across domains. While we examined a broad range of pre-specified primary and secondary outcomes reflecting the integrated content areas covered by the intervention, this breadth also poses challenges for interpreting overall intervention effects across domains with multiple outcome measures, particularly when effects vary in magnitude or direction across individual indicators.

These domain-level summary indices aggregate multiple related outcomes into a single summary measure, while mitigating the influence of measurement errors in individual indicators and capturing broader underlying constructs targeted by the intervention. These domain-level summary tests are particularly useful for evaluating comprehensive, multi-component interventions such as the *Healthy Future* program with integrated content across multiple dimensions of maternal and child health.

**Table A.** Comparison of two intervention arms.

|  | **Standard**<br>**Arm**<br>**(n=278)** | **Family Encouragement**<br>**Arm**<br>**(n=284)** | **Difference**<br>**P-value** |
| --- | --- | --- | --- |
| **Intervention take-up** |  |  |  |
| Number of families completed at least one home visit (0/1) | 231 (83%) | 230 (81%) | 0.52 |
| **Family participation in completed home visits** |  |  |  |
| % of primary caregiver participated, mean (SD) | 0.955 (0.106) | 0.967 (0.072) | 0.17 |
| % of secondary caregiver participated, mean (SD) | 0.213 (0.251) | 0.450 (0.315) | <0.001 |

**Table B.**
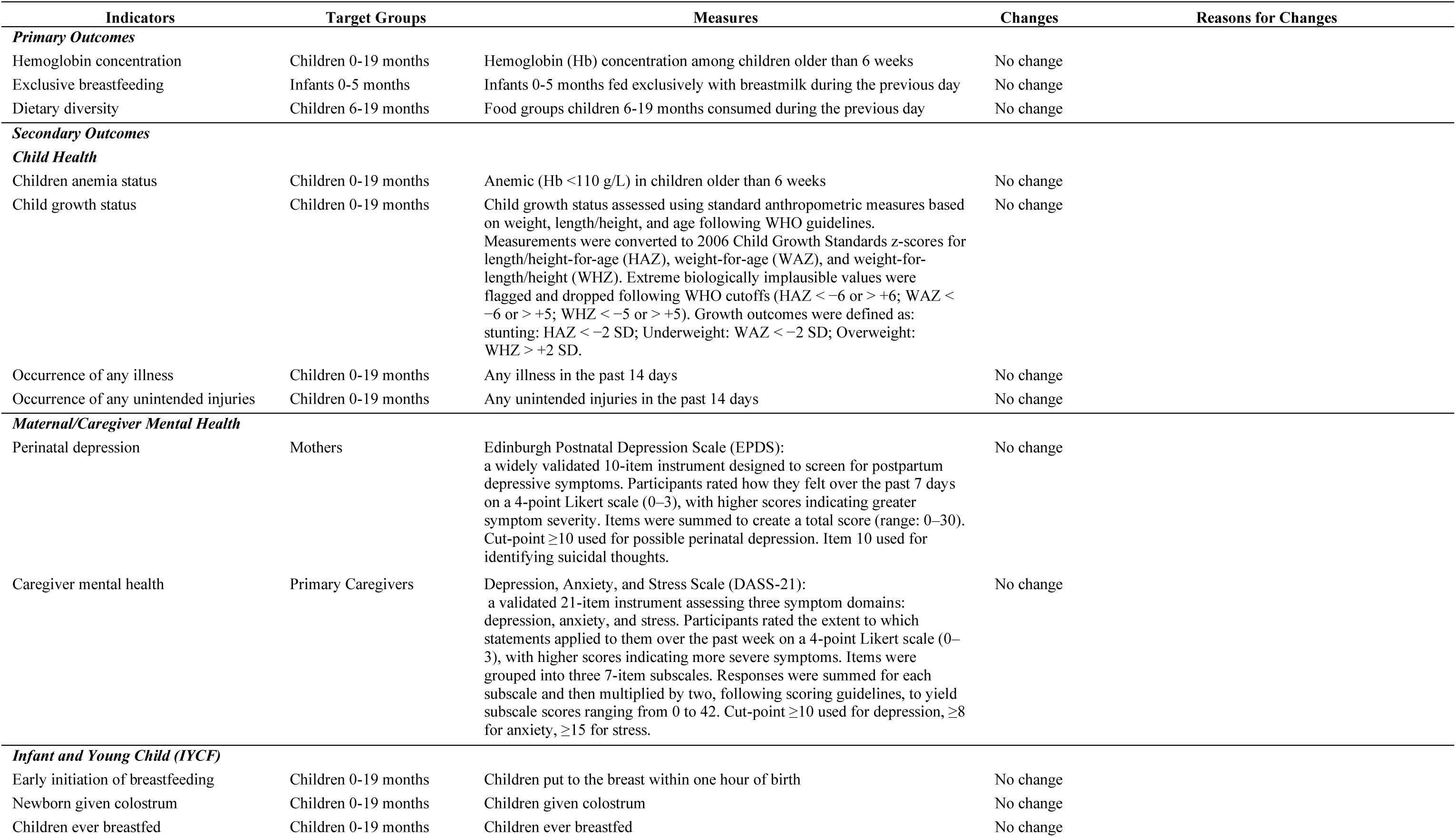
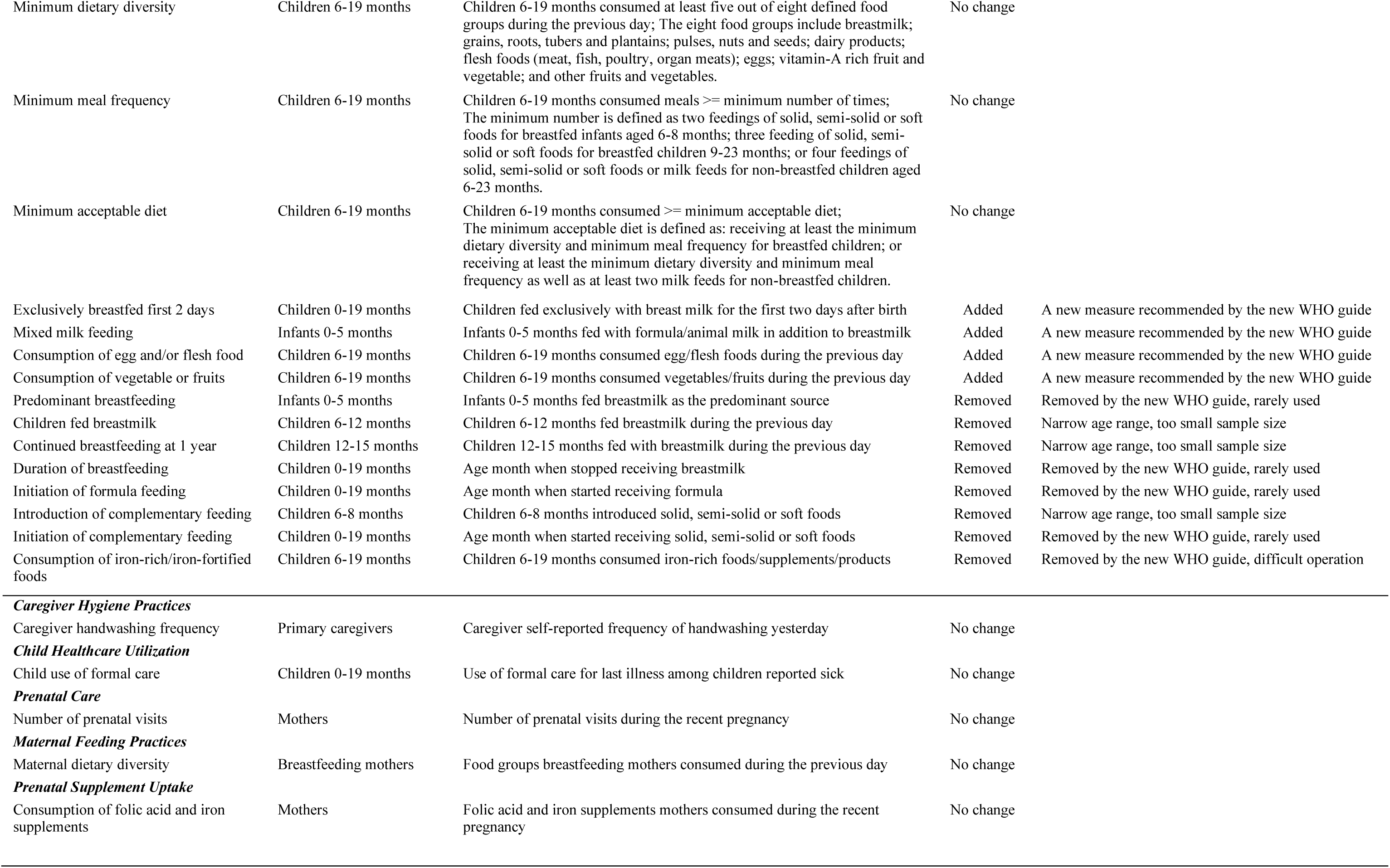
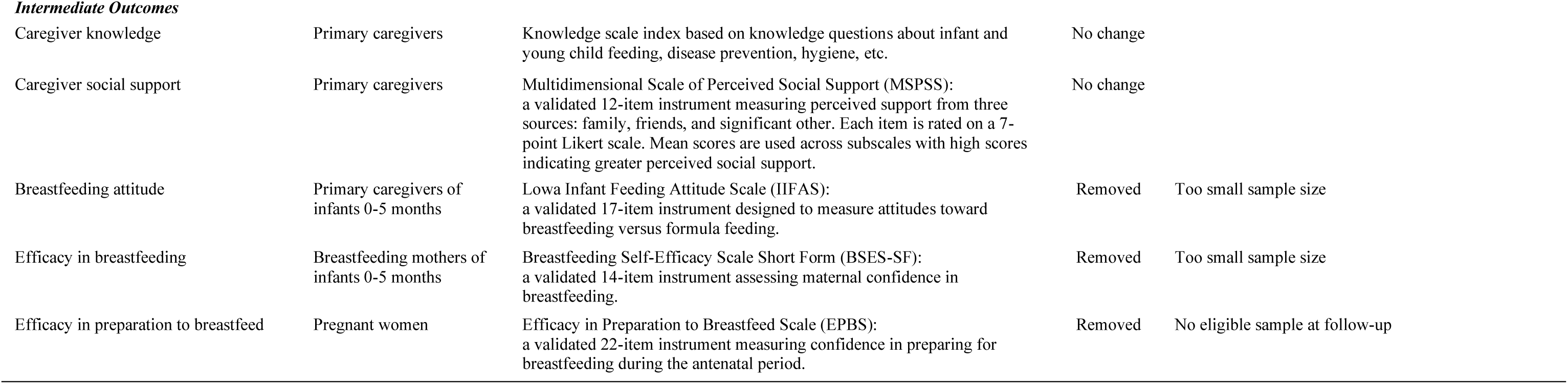
Study outcome measures and changes from the protocol.

| Indicators | Target Groups | Measures | Changes | Reasons for Changes |
| --- | --- | --- | --- | --- |
| **Primary Outcomes** |  |  |  |  |
| Hemoglobin concentration | Children 0-19 months | Hemoglobin (Hb) concentration among children older than 6 weeks | No change |  |
| Exclusive breastfeeding | Infants 0-5 months | Infants 0-5 months fed exclusively with breastmilk during the previous day | No change |  |
| Dietary diversity | Children 6-19 months | Food groups children 6-19 months consumed during the previous day | No change |  |
| **Secondary Outcomes** |  |  |  |  |
| **Child Health** |  |  |  |  |
| Children anemia status | Children 0-19 months | Anemic (Hb <110 g/L) in children older than 6 weeks | No change |  |
| Child growth status | Children 0-19 months | Child growth status assessed using standard anthropometric measures based on weight, length/height, and age following WHO guidelines. Measurements were converted to 2006 Child Growth Standards z-scores for length/height-for-age (HAZ), weight-for-age (WAZ), and weight-for-length/height (WHZ). Extreme biologically implausible values were flagged and dropped following WHO cutoffs (HAZ < -6 or > +6; WAZ < -6 or > +5; WHZ < -5 or > +5). Growth outcomes were defined as: stunting: HAZ < -2 SD; Underweight: WAZ < -2 SD; Overweight: WHZ > +2 SD. | No change |  |
| Occurrence of any illness | Children 0-19 months | Any illness in the past 14 days | No change |  |
| Occurrence of any unintended injuries | Children 0-19 months | Any unintended injuries in the past 14 days | No change |  |
| **Maternal/Caregiver Mental Health** |  |  |  |  |
| Perinatal depression | Mothers | Edinburgh Postnatal Depression Scale (EPDS): a widely validated 10-item instrument designed to screen for postpartum depressive symptoms. Participants rated how they felt over the past 7 days on a 4-point Likert scale (0–3), with higher scores indicating greater symptom severity. Items were summed to create a total score (range: 0–30). Cut-point ≥10 used for possible perinatal depression. Item 10 used for identifying suicidal thoughts. | No change |  |
| Caregiver mental health | Primary Caregivers | Depression, Anxiety, and Stress Scale (DASS-21): a validated 21-item instrument assessing three symptom domains: depression, anxiety, and stress. Participants rated the extent to which statements applied to them over the past week on a 4-point Likert scale (0–3), with higher scores indicating more severe symptoms. Items were grouped into three 7-item subscales. Responses were summed for each subscale and then multiplied by two, following scoring guidelines, to yield subscale scores ranging from 0 to 42. Cut-point ≥10 used for depression, ≥8 for anxiety, ≥15 for stress. | No change |  |
| **Infant and Young Child (IYCF)** |  |  |  |  |
| Early initiation of breastfeeding | Children 0-19 months | Children put to the breast within one hour of birth | No change |  |
| Newborn given colostrum | Children 0-19 months | Children given colostrum | No change |  |
| Children ever breastfed | Children 0-19 months | Children ever breastfed | No change |  |
| Minimum dietary diversity | Children 6-19 months | Children 6-19 months consumed at least five out of eight defined food groups during the previous day; The eight food groups include breastmilk; grains, roots, tubers and plantains; pulses, nuts and seeds; dairy products; flesh foods (meat, fish, poultry, organ meats); eggs; vitamin-A rich fruit and vegetable; and other fruits and vegetables. | No change |  |
| Minimum meal frequency | Children 6-19 months | Children 6-19 months consumed meals ≥ minimum number of times; The minimum number is defined as two feedings of solid, semi-solid or soft foods for breastfed infants aged 6-8 months; three feeding of solid, semi-solid or soft foods for breastfed children 9-23 months; or four feedings of solid, semi-solid or soft foods or milk feeds for non-breastfed children aged 6-23 months. | No change | |
| Minimum acceptable diet | Children 6-19 months | Children 6-19 months consumed ≥ minimum acceptable diet; The minimum acceptable diet is defined as: receiving at least the minimum dietary diversity and minimum meal frequency for breastfed children; or receiving at least the minimum dietary diversity and minimum meal frequency as well as at least two milk feeds for non-breastfed children. | No change | |
| Exclusively breastfed first 2 days | Children 0-19 months | Children fed exclusively with breast milk for the first two days after birth | Added | A new measure recommended by the new WHO guide |
| Mixed milk feeding | Infants 0-5 months | Infants 0-5 months fed with formula/animal milk in addition to breastmilk | Added | A new measure recommended by the new WHO guide |
| Consumption of egg and/or flesh food | Children 6-19 months | Children 6-19 months consumed egg/flesh foods during the previous day | Added | A new measure recommended by the new WHO guide |
| Consumption of vegetable or fruits | Children 6-19 months | Children 6-19 months consumed vegetables/fruits during the previous day | Added | A new measure recommended by the new WHO guide |
| Predominant breastfeeding | Infants 0-5 months | Infants 0-5 months fed breastmilk as the predominant source | Removed | Removed by the new WHO guide, rarely used |
| Children fed breastmilk | Children 6-12 months | Children 6-12 months fed breastmilk during the previous day | Removed | Narrow age range, too small sample size |
| Continued breastfeeding at 1 year | Children 12-15 months | Children 12-15 months fed with breastmilk during the previous day | Removed | Narrow age range, too small sample size |
| Duration of breastfeeding | Children 0-19 months | Age month when stopped receiving breastmilk | Removed | Removed by the new WHO guide, rarely used |
| Initiation of formula feeding | Children 0-19 months | Age month when started receiving formula | Removed | Removed by the new WHO guide, rarely used |
| Introduction of complementary feeding | Children 6-8 months | Children 6-8 months introduced solid, semi-solid or soft foods | Removed | Narrow age range, too small sample size |
| Initiation of complementary feeding | Children 0-19 months | Age month when started receiving solid, semi-solid or soft foods | Removed | Removed by the new WHO guide, rarely used |
| Consumption of iron-rich/iron-fortified foods | Children 6-19 months | Children 6-19 months consumed iron-rich foods/supplements/products | Removed | Removed by the new WHO guide, difficult operation |
| **Caregiver Hygiene Practices** |  |  |  |  |
| Caregiver handwashing frequency | Primary caregivers | Caregiver self-reported frequency of handwashing yesterday | No change |  |
| **Child Healthcare Utilization** |  |  |  |  |
| Child use of formal care | Children 0-19 months | Use of formal care for last illness among children reported sick | No change |  |
| **Prenatal Care** |  |  |  |  |
| Number of prenatal visits | Mothers | Number of prenatal visits during the recent pregnancy | No change |  |
| **Maternal Feeding Practices** |  |  |  |  |
| Maternal dietary diversity | Breastfeeding mothers | Food groups breastfeeding mothers consumed during the previous day | No change |  |
| **Prenatal Supplement Uptake** |  |  |  |  |
| Consumption of folic acid and iron supplements | Mothers | Folic acid and iron supplements mothers consumed during the recent pregnancy | No change |  |
| ***Intermediate Outcomes*** |  |  |  |  |
| Caregiver knowledge | Primary caregivers | Knowledge scale index based on knowledge questions about infant and young child feeding, disease prevention, hygiene, etc. | No change |  |
| Caregiver social support | Primary caregivers | Multidimensional Scale of Perceived Social Support (MSPSS): a validated 12-item instrument measuring perceived support from three sources: family, friends, and significant other. Each item is rated on a 7-point Likert scale. Mean scores are used across subscales with high scores indicating greater perceived social support. | No change |  |
| Breastfeeding attitude | Primary caregivers of infants 0-5 months | Lowa Infant Feeding Attitude Scale (IIFAS): a validated 17-item instrument designed to measure attitudes toward breastfeeding versus formula feeding. | Removed | Too small sample size |
| Efficacy in breastfeeding | Breastfeeding mothers of infants 0-5 months | Breastfeeding Self-Efficacy Scale Short Form (BSES-SF): a validated 14-item instrument assessing maternal confidence in breastfeeding. | Removed | Too small sample size |
| Efficacy in preparation to breastfeed | Pregnant women | Efficacy in Preparation to Breastfeed Scale (EPBS): a validated 22-item instrument measuring confidence in preparing for breastfeeding during the antenatal period. | Removed | No eligible sample at follow-up |

### S3. Statistical Analysis Details

#### 1. Intent-to-treat (ITT) specification

The main evaluation employed ITT analysis to estimate the effects of offering the program to eligible families. We regressed the outcomes of interest at the follow-up surveys on a treatment indicator on township assignment, baseline controls, county stratum fixed effects using the following specification:

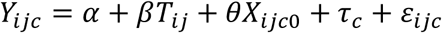

*Y*_*ijc*_ is the outcome of interest for individual *i* (child or caregiver) in township *j* of county *c* measured at follow-up; *T*_*ij*_ is an indicator for whether the individual *i* in township *j* was assigned to the treatment arm; *τ*_*c*_ denotes randomization strata (i.e., county *c*) fixed effects; *X*_*ijc*0_ is a vector of baseline control variables, capturing child, caregiver, and household characteristics measured at baseline, selected using a machine-learning post-double-selection lasso procedure. Standard errors were clustered at the township level using the cluster-corrected Huber-White estimator.

The treatment indicator, *T*_*j*_, takes the value one if the family lives in the treatment township. This approach identifies the intent-to-treat effect of offering the program, taking into account that some families in the treatment group may not participate in the program. The main parameter of interest, *β*, represents the Average Treatment Effects and is interpreted as the causal effects of offering the program.

#### 2. Compliance-adjusted treatment-on-the-treated (TOT) specification

Because participation in the program was voluntary, not all families assigned to intervention townships received the same level of intervention exposure. To account for imperfect compliance, we conducted a compliance-adjusted treatment-on-the-treated (TOT) analysis using an instrumental variable (IV) approach.

In this specification, intervention exposure was measured as the cumulative number of completed home visits prior to the follow-up survey, with control families assigned a value of zero. Because the number of completed home visits may be correlated with unobserved family characteristics that also affect outcomes, direct estimation of the association between home visits and outcomes may be subject to endogeneity bias. We therefore used assignment to an intervention township as an instrument for the cumulative number of completed home visits. Random assignment ensures that treatment assignment is exogenous, while assignment to an intervention township strongly predicts intervention exposure through participation in the program.

Under the standard IV assumptions, this approach identifies the causal effect of additional intervention exposure induced by assignment to the intervention group. The resulting estimates can be interpreted as compliance-adjusted treatment effects on the treated among participating families whose participation was influenced by treatment assignment.

#### 3. Selection of control variables using the post-double-selection lasso procedure

To improve the precision of treatment effect estimates, we selected baseline control variables using the post-double-selection lasso procedure proposed by Belloni, Chernozhukov, and Hansen (2014). In randomized trials, treatment assignment is independent of baseline characteristics by design. However, including strong predictors of the outcome can substantially improve statistical precision and increase power. Because the appropriate set of covariates is often unknown ex ante, the post-double-selection lasso procedure provides a data-driven approach for selecting relevant control variables from a large set of baseline characteristics while limiting overfitting.

The procedure identifies appropriate control variables through two separate lasso regressions. In the first step, baseline variables that predict the outcome are selected. In the second step, baseline variables that predict the treatment variable are selected. Variables identified in either step are then included in the final regression model. This approach helps ensure that important predictors of either the outcome or treatment assignment are retained in the estimation.

Potential control variables were drawn from a broad set of baseline characteristics measured at the child, caregiver, and household levels (Table A). We followed the standard cleaning procedure for all control variables: continuous variables were standardized, categorical variables were converted into indicator variables, and squared terms were generated for all continuous variables to allow for potential nonlinear relationships. For variables with missing values, we created missing-value indicators and imputed using township-specific means. Randomization strata fixed effects were partialled out prior to lasso estimation to ensure that stratification variables were always included in the analysis.

This procedure was applied to both the ITT and TOT analyses. ITT estimates were obtained using the *pdslasso* command, while TOT estimates were obtained using the *ivlasso* command, from the Stata package *lassopack* developed by Ahrens, Hansen, and Schaffer (2020).

#### 4. Multiple inference adjustments for secondary outcomes

Because the study evaluates a large number of secondary outcomes across multiple domains, we adjusted for multiple hypothesis testing when interpreting secondary outcome results.

Specifically, we controlled the false discovery rate (FDR), defined as the expected proportion of rejected hypotheses that are false positives. FDR control is commonly used when evaluating multiple related outcomes because it balances protection against Type-I error with statistical power and is often considered more appropriate than family-wise error rate adjustments for exploratory analyses involving numerous secondary outcomes.

Because the *Healthy Future* program targeted multiple distinct domains, we treated each domain as a separate family of related hypotheses and applied FDR adjustment within domains. We calculated FDR-adjusted p-values within each outcome domain using the two-stage procedure proposed by Benjamini, Krieger, and Yekutieli (2006). This procedure estimates the proportion of true null hypotheses and provides sharpened control of the false discovery rate. The use of domain-specific FDR adjustment follows the approach recommended by Anderson (2008) for studies evaluating multiple related outcomes.

#### 5. Construction of domain-level summary indices

We also constructed domain-level summary indices that aggregated primary and secondary outcomes within conceptually related domains. These analyses were not pre-specified in the study protocol and were conducted as supplementary analyses. Because the *Healthy Future* program targeted multiple domains, these domain-level summary indices provide high-level interpretations of the overall interventional effects across domains.

We followed the generalized least squares (GLS) weighting procedure proposed by Anderson (2008). This approach constructs a summary index as a weighted average of multiple standardized outcomes within a domain. This weighting procedure accounts for the correlation structure among outcomes, assigning relatively smaller weights to highly correlated measures and larger weights to measures that contribute additional independent information. As a result, the index provides a single summary measure of intervention effects across multiple related outcomes within a domain.

The following steps were taken from Anderson (2008). The procedure was implemented using the *swindex* command in Stata:

1. All outcomes were coded so that higher values consistently represented better outcomes.
2. Outcomes were standardized by subtracting the control-group mean and dividing by the control-group standard deviation.
3. Outcomes were grouped into conceptually related domains, with each outcome assigned to a single domain.
4. For each domain, a summary index was constructed as a weighted average of the standardized outcomes using weights derived from the inverse covariance matrix of the outcomes within that domain.
5. The resulting domain-level summary indices were analyzed using the same regression specifications as the individual outcomes to estimate intervention effects at the domain level.

Table B provides the list of primary and secondary outcome measures included in constructing each domain-level summary index. The same index construction procedure was applied consistently across all domains to ensure comparability of intervention effect estimates.

**Table A.**
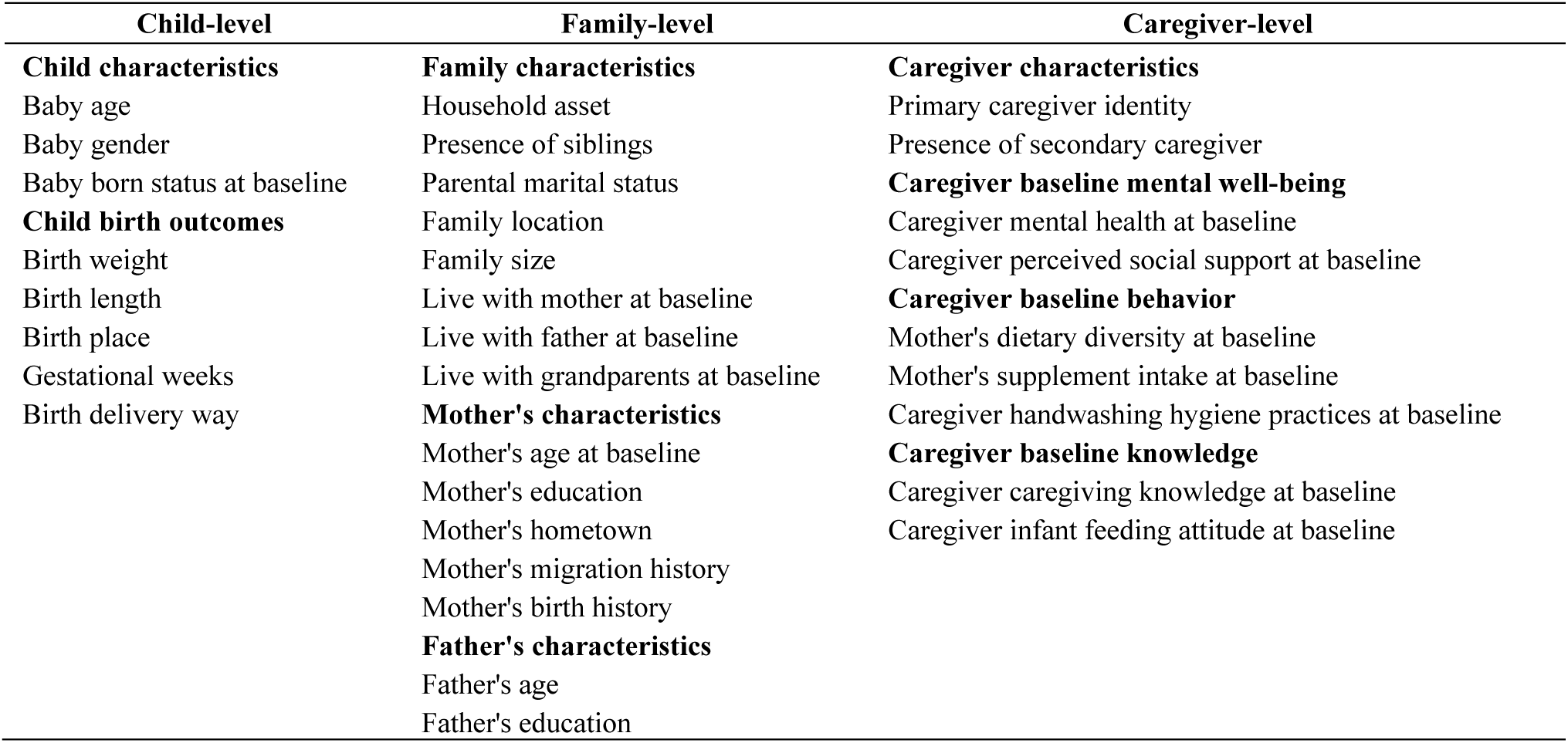
List of control variables used in the post-double-selection lasso procedure.

**Table B.** List of individual measures for constructing domain-level summary indices.

| Domain Summary Index | Individual Measures Included | Primary or Secondary Outcome |
| --- | --- | --- |
| *Child-level Outcomes* |  |  |
| Child health index (children) | Hemoglobin level (g/L) | Primary |
|  | Anemia (0/1) | Secondary |
|  | Stunted (0/1) | Secondary |
|  | Underweight (0/1) | Secondary |
|  | Overweight (0/1) | Secondary |
|  | Any illness in the past 14 days (0/1) | Secondary |
|  | Any unintended injuries in the past 14 days (0/1) | Secondary |
| Breastfeeding initiation index (children) | Early initiation of breastfeeding (0/1) | Secondary |
|  | Newborn given colostrum (0/1) | Secondary |
|  | Children ever breastfed (0/1) | Secondary |
|  | Exclusively breastfed first 2 days (0/1) | Secondary |
| Breastfeeding index (infants 0-5 months) | Exclusive breastfeeding (0/1) | Primary |
|  | Mixed milk feeding (0/1) | Secondary |
| Complementary feeding index (children 6-19 months) | Dietary diversity score (food groups) | Primary |
|  | Minimum dietary diversity (0/1) | Secondary |
|  | Minimum meal frequency (0/1) | Secondary |
|  | Minimum acceptable diet (0/1) | Secondary |
|  | Consumption of egg and/or flesh food (0/1) | Secondary |
|  | Consumption of vegetable or fruits (0/1) | Secondary |
| Child caregiving knowledge index (primary caregivers) | Child caregiving knowledge index constructed based on caregiver knowledge questions on infant and young child feeding, disease prevention, hygiene, etc. | Secondary |
| *Mother/Caregiver-level Outcomes* |  |  |
| Mental health index (mothers) | Possible perinatal depression (0/1) | Secondary |
|  | Suicidal thoughts (0/1) | Secondary |
|  | EPDS score | Secondary |
| Mental health index (primary caregivers) | Depression (0/1) | Secondary |
|  | Anxiety (0/1) | Secondary |
|  | Stress (0/1) | Secondary |
|  | DASS-21 score | Secondary |
| Mental health self-management knowledge index (primary caregivers) | Caregiver mental health self-management knowledge index based on questions on mental health self-management | Secondary |
| Perceived social support index (primary caregivers) | Perceived support from family (score) | Secondary |
|  | Perceived support from friends (score) | Secondary |
|  | Perceived support from significant other (score) | Secondary |

## S4. Results from the Compliance-Adjusted Treatment-on-the-Treated (TOT) Analysis

**Table A.**
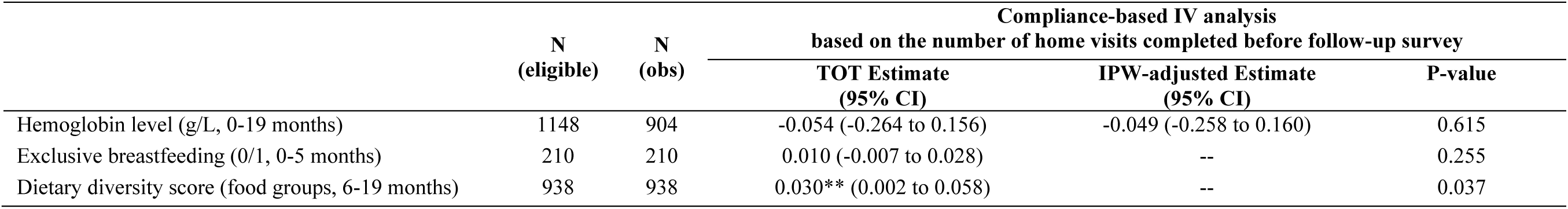
TOT effects on primary outcomes.

**Table B.** TOT effects on secondary health outcomes.

|  | N<br>(eligible) | N<br>(obs) | Control<br>Mean | Compliance-based IV analysis |  |  |  |
| --- | --- | --- | --- | --- | --- | --- | --- |
|  |  |  |  | based on the number of home visits completed before follow-up survey |  |  |  |
|  |  |  |  | TOT Estimate<br>(95% CI) | IPW-adjusted Estimate<br>(95% CI) | P-value | FDR-adjusted<br>P-value |
| **Child Health (Children)** |  |  |  |  |  |  |  |
| Anemia (0/1) | 1148 | 904 | 0.421 | 0.005 (-0.003 to 0.014) | 0.005 (-0.003 to 0.014) | 0.224 | 1.000 |
| Stunted (0/1) | 1148 | 959 | 0.061 | -0.002 (-0.005 to 0.002) | -0.001 (-0.005 to 0.002) | 0.450 | 1.000 |
| Underweight (0/1) | 1148 | 951 | 0.014 | -0.001 (-0.002 to 0.001) | -0.001 (-0.003 to 0.001) | 0.526 | 1.000 |
| Overweight (0/1) | 1148 | 946 | 0.152 | -0.002 (-0.008 to 0.004) | -0.002 (-0.008 to 0.004) | 0.463 | 1.000 |
| Any illness (0/1) | 1148 | 1148 | 0.643 | -0.012*** (-0.020 to -0.003) | -- | 0.007 | 0.045 |
| Any unintended injuries (0/1) | 1148 | 1148 | 0.092 | 0.001 (-0.004 to 0.006) | -- | 0.655 | 1.000 |
| **Perinatal Mental Health**<br>**(Mothers)** |  |  |  |  |  |  |  |
| Possible perinatal depression (0/1) | 1148 | 827 | 0.178 | -0.007** (-0.013 to -0.001) | -0.008** (-0.015 to -0.001) | 0.020 | 0.031 |
| Suicidal thoughts (0/1) | 1148 | 827 | 0.056 | -0.001 (-0.005 to 0.003) | -0.002 (-0.006 to 0.003) | 0.538 | 0.219 |
| EPDS score | 1148 | 827 | 5.212 | -0.105** (-0.187 to -0.023) | -0.123*** (-0.210 to -0.036) | 0.012 | 0.031 |
| **Caregiver Mental Health**<br>**(Primary Caregivers)** |  |  |  |  |  |  |  |
| Depression (0/1) | 1148 | 973 | 0.169 | -0.009*** (-0.015 to -0.002) | -0.008*** (-0.015 to -0.002) | 0.006 | 0.025 |
| Anxiety (0/1) | 1148 | 973 | 0.171 | -0.007** (-0.012 to -0.001) | -0.006** (-0.012 to -0.001) | 0.017 | 0.027 |
| Stress (0/1) | 1148 | 973 | 0.120 | -0.006** (-0.011 to -0.000) | -0.005** (-0.011 to -0.000) | 0.037 | 0.029 |
| DASS-21 score | 1148 | 973 | 13.598 | -0.323** (-0.623 to -0.023) | -0.315** (-0.615 to -0.014) | 0.035 | 0.029 |

**Table C.** TOT effects on secondary infant and young child feeding outcomes.

|  | N<br>(eligible) | N<br>(obs) | Control<br>Mean | Compliance-based IV analysis<br>based on the number of home visits completed before follow-up survey |  |  |
| --- | --- | --- | --- | --- | --- | --- |
|  |  |  |  | TOT Estimate<br>(95% CI) | P-value | FDR-adjusted<br>P-value |
| **Breastfeeding Initiation (All children)** |  |  |  |  |  |  |
| Early initiation of breastfeeding (0/1) | 1148 | 1148 | 0.189 | 0.007 (-0.000 to 0.014) | 0.055 | 0.058 |
| Newborn given colostrum (0/1) | 1148 | 1148 | 0.824 | 0.006** (0.000 to 0.013) | 0.042 | 0.058 |
| Children ever breastfed (0/1) | 1148 | 1148 | 0.937 | 0.005*** (0.002 to 0.008) | 0.002 | 0.009 |
| Exclusively breastfed first 2 days (0/1) | 1148 | 1148 | 0.100 | 0.002 (-0.004 to 0.008) | 0.481 | 0.137 |
| **Breastfeeding Practice (Infants <6 months)** |  |  |  |  |  |  |
| Mixed milk feeding (0/1) | 210 | 210 | 0.203 | -0.007 (-0.021 to 0.006) | 0.285 | -- |
| **Complementary Feeding (Children >6 months)** |  |  |  |  |  |  |
| Minimum dietary diversity (0/1) | 938 | 938 | 0.505 | 0.006 (-0.003 to 0.014) | 0.171 | 0.148 |
| Minimum meal frequency (0/1) | 938 | 938 | 0.692 | 0.004 (-0.003 to 0.012) | 0.272 | 0.195 |
| Minimum acceptable diet (0/1) | 938 | 938 | 0.381 | 0.006 (-0.002 to 0.014) | 0.139 | 0.148 |
| Consumption of egg and/or flesh food (0/1) | 938 | 938 | 0.624 | 0.008** (0.001 to 0.016) | 0.025 | 0.066 |
| Consumption of vegetable or fruits (0/1) | 938 | 938 | 0.763 | 0.008** (0.001 to 0.014) | 0.017 | 0.066 |

**Table D.** TOT effects on hygiene, healthcare utilization, and prenatal care.

|  | N<br>(eligible) | N<br>(obs) | Control<br>Mean | Compliance-based IV analysis<br>based on the number of home visits completed before follow-up |  |  |
| --- | --- | --- | --- | --- | --- | --- |
|  |  |  |  | TOT Estimate<br>(95% CI) | IPW-adjusted Estimate<br>(95% CI) | P-value |
| **Caregiver Hygiene Practices (Primary Caregivers)** |  |  |  |  |  |  |
| Caregiver handwashing frequency (number) | 1148 | 1121 | 5.509 | 0.018 (-0.054 to 0.089) | 0.118 (-0.054 to 0.089) | 0.625 |
| **Child Healthcare Utilization (Children)** |  |  |  |  |  |  |
| Use of formal care for last illness (0/1) | 1148 | 950 | 0.834 | 0.000 (-0.006 to 0.007) | 0.000 (-0.006 to 0.007) | 0.968 |
| **Prenatal Care (Mothers)** |  |  |  |  |  |  |
| Prenatal visits during recent pregnancy (number) | 1148 | 1085 | 4.261 | 0.007 (-0.017 to 0.031) | 0.007 (-0.017 to 0.031) | 0.563 |
| **Maternal Feeding (Mothers Currently Breastfeeding)** |  |  |  |  |  |  |
| Maternal dietary diversity (food groups) | n/a | 449 | 5.797 | 0.026 (-0.008 to 0.061) | -- | 0.136 |
| **Prenatal Supplement Uptake (Mothers)** |  |  |  |  |  |  |
| Consumption of folic acid and iron supplements during pregnancy (0/1) | 1148 | 1089 | 0.576 | -0.000 (-0.006 to 0.006) | -0.000 (-0.006 to 0.006) | 0.973 |

**Table E.**
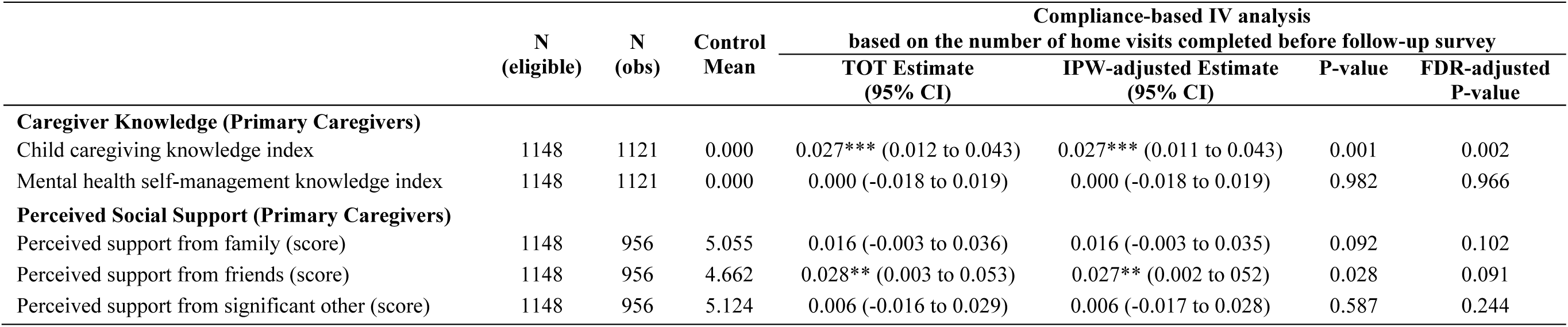
TOT effects on intermediate outcomes.

|  | N<br>(eligible) | N<br>(obs) | Control<br>Mean | Compliance-based IV analysis |  |  |  |
| --- | --- | --- | --- | --- | --- | --- | --- |
|  |  |  |  | based on the number of home visits completed before follow-up survey |  |  |  |
|  |  |  |  | TOT Estimate<br>(95% CI) | IPW-adjusted Estimate<br>(95% CI) | P-value | FDR-adjusted<br>P-value |
| **Caregiver Knowledge (Primary Caregivers)** |  |  |  |  |  |  |  |
| Child caregiving knowledge index | 1148 | 1121 | 0.000 | 0.027*** (0.012 to 0.043) | 0.027*** (0.011 to 0.043) | 0.001 | 0.002 |
| Mental health self-management knowledge index | 1148 | 1121 | 0.000 | 0.000 (-0.018 to 0.019) | 0.000 (-0.018 to 0.019) | 0.982 | 0.966 |
| **Perceived Social Support (Primary Caregivers)** |  |  |  |  |  |  |  |
| Perceived support from family (score) | 1148 | 956 | 5.055 | 0.016 (-0.003 to 0.036) | 0.016 (-0.003 to 0.035) | 0.092 | 0.102 |
| Perceived support from friends (score) | 1148 | 956 | 4.662 | 0.028** (0.003 to 0.053) | 0.027** (0.002 to 0.052) | 0.028 | 0.091 |
| Perceived support from significant other (score) | 1148 | 956 | 5.124 | 0.006 (-0.016 to 0.029) | 0.006 (-0.017 to 0.028) | 0.587 | 0.244 |

**Table F.** TOT effects on domain-level summary indices.

|  | N<br>(eligible) | N<br>(obs) | Control<br>Mean | Compliance-based IV analysis<br>based on the number of home visits completed before follow-up |  |  |
| --- | --- | --- | --- | --- | --- | --- |
|  |  |  |  | TOT Estimate<br>(95% CI) | IPW-adjusted Estimate<br>(95% CI) | P-value |
| Child-level Outcomes |  |  |  |  |  |  |
| Child health index (children) | 1148 | 1148 | 0.000 | 0.011 (-0.005 to 0.028) | -- | 0.175 |
| Breastfeeding initiation index (children) | 1148 | 1148 | 0.000 | 0.024*** (0.008 to 0.040) | -- | 0.004 |
| Breastfeeding index (infants 0-5 months) | 210 | 210 | 0.000 | 0.030 (-0.006 to 0.066) | -- | 0.106 |
| Complementary feeding index (children 6-19 months) | 938 | 938 | 0.000 | 0.018*** (0.004 to 0.032) | -- | 0.010 |
| Caregiving knowledge index (primary caregivers) | 1148 | 1121 | 0.000 | 0.027*** (0.012 to 0.043) | 0.027*** (0.011 to 0.043) | 0.001 |
| Mother/Caregiver-level Outcomes |  |  |  |  |  |  |
| Mental health index (mothers) | 1148 | 827 | 0.000 | 0.017** (0.001 to 0.033) | 0.020** (0.002 to 0.039) | 0.038 |
| Mental health index (primary caregivers) | 1148 | 973 | 0.000 | 0.022*** (0.007 to 0.037) | 0.021*** (0.006 to 0.036) | 0.004 |
| Mental health self-management knowledge (primary caregivers) | 1148 | 1121 | 0.000 | 0.000 (-0.018 to 0.019) | 0.000 (-0.018 to 0.019) | 0.982 |
| Perceived social support index (primary caregivers) | 1148 | 956 | 0.000 | 0.016** (0.001 to 0.031) | 0.116** (0.001 to 0.031) | 0.032 |

